# Left-handedness, learning disability, autoimmune disease, and seizure history influence age at onset and phenotypical targeting of Alzheimer’s disease

**DOI:** 10.1101/2022.12.17.22283307

**Authors:** Zachary A. Miller, Rik Ossenkoppele, Neill R. Graff-Radford, Isabel E. Allen, Wendy Shwe, Lynne Rosenberg, Dustin J Olguin, Michael G. Erkkinen, P. Monroe Butler, Salvatore Spina, Jennifer S. Yokoyama, Rahul S. Desikan, Philip Scheltens, Wiesje van der Flier, Yolande Pijnenburg, Emma Wolters, Rosa Rademakers, Daniel H. Geschwind, Joel H. Kramer, Howard J. Rosen, Katherine P. Rankin, Lea T. Grinberg, William W. Seeley, Virginia Sturm, David C. Perry, Bruce L. Miller, Gil D. Rabinovici, Maria Luisa Gorno-Tempini

**Author notes:** **Correspondence:** Zachary A. Miller, UCSF Memory and Aging Center, 675 Nelson Rising Lane, Suite 191D Box 1207 San Francisco, CA 94143-1207, USA. Dr. Rahul Desikan passed away on July 14^th^, 2019. These authors contributed equally to this work and share last authorship (Bruce L. Miller, Gil D. Rabinovici, Maria Luisa Gorno-Tempini).

## Abstract

**Background:** Risk factors associated with sporadic non-amnestic and early-onset Alzheimer’s disease remain underexamined. We investigated a large, clinically heterogeneous Alzheimer’s disease cohort for frequencies of established Alzheimer’s disease risk factors (hypertension, hyperlipidemia, diabetes mellitus, *APOE*-ɛ4 frequency, and years of education), alongside a suite of novel factors with historical theoretical association (non-right-handedness, learning disability, seizures, and autoimmune disease).

**Methods:** In this case-control study, we screened the demographic and health histories of 750 consecutive early-onset and 750 late-onset Alzheimer’s disease patients from the University of California San Francisco Memory and Aging Center for the prevalence of conventional risk and novel Alzheimer’s disease factors and compared these results with 8,859 Alzheimer’s disease individuals from the National Alzheimer’s Coordinating Center, Amsterdam University Medical Center, Amsterdam, and Mayo Clinic, Jacksonville.

**Results:** Early-onset Alzheimer’s disease was associated with significantly lower frequencies of established risk factors (hypertension, hyperlipidemia, diabetes mellitus, all *p*<0.001, *APOE*-ɛ4, *p*=0.03) and significantly higher frequencies of novel factors (non-right-handedness, learning disability, active seizure, all *p*<0.001, remote seizure, *p*=0.002, and autoimmune disease, *p*=0.007). Logistic regressions predicting EOAD vs. LOAD controlling for sex, education, *APOE*-ɛ4 status, typical, and novel risk factors, produced findings consistent with the above. Principal component analysis loaded novel factors into two components, non-right-handedness and learning disability versus seizure and autoimmune disease, and the combination of factors from both components resulted in an exponential decrease in age at onset from any single factor alone. *APOE*-ɛ4 provided no additional contribution to age at onset decreases within the non-amnestic Alzheimer’s disease cohort but shifted the age of onset 3 years earlier within amnestic presentations (*p*=0.013).

**Conclusions:** We identified non-right-handedness, learning disability, seizures, and autoimmune disease as novel factors that affect both the age at onset and phenotypical targeting of Alzheimer’s disease. Together these results support a new theoretical framework of neurodegenerative disease susceptibility and that through the collection of detailed developmental and health history, neurodegenerative disease risk in some may be highly predictable, offering new opportunities towards early detection, monitoring, therapeutic intervention, and ultimately disease prevention.

## Background

Alzheimer’s disease (AD) is the largest contributor to dementia world-wide. With a rapidly aging population, lacking powerful interventions, the world faces a healthcare crisis. To date, mitigating the effects of established AD risk factors like hypertension, hypercholesterolemia, diabetes mellitus, and limited schooling, through advancements in the treatment of vascular disease and improved access to education, is beginning to have an impact on reducing the global burden of AD.(1) As established AD associated risk factors come almost exclusively from studies of typical amnestic late-onset AD (LOAD) (age at first symptoms of ≥65) and are largely age-related, we sought to identify novel AD associated risk factors by examining the demographic and health histories of individuals presenting with early-onset AD (EOAD).

The distinction between EOAD and LOAD is historic and largely arbitrary, based on the misconception that declines in cognitive status at or after age 65 reflected a distinctly different process than declines witnessed prior to age 65.(2) Nevertheless, meaningful differences between early and late presentations of AD exist. EOAD is less common than LOAD, accounting for 4-10% of all AD. Typically, AD is a memory-predominant, hippocampal-based amnestic syndrome, but EOAD cases demonstrate a greater variety of clinical presentations that include: behavioral, dysexecutive, apraxic, visuospatial, or focal language AD clinical presentations. Together these variants are referred to as atypical focal cortical or non-amnestic AD. Non-amnestic AD presentations are also more common within EOAD, constituting up to 25-50% of EOAD compared to <10% of LOAD presentations. Furthermore, the most common genetic risk factor for AD, *APOE*-ɛ4, is less common in EOAD, most notably so in non-amnestic AD presentations.(3) Despite sharing the same underlying pathological substrate as LOAD, the reasons for the earlier age at onset, greater degree of clinical heterogeneity, and decreased frequency of *APOE*-ɛ4 in EOAD remains poorly understood.

Recognizing that neurodegenerative disorders start focally, spread in a network-based fashion, and produce distinctive clinical phenotypes,(4) we identified that neurodevelopmental differences (non-right-handedness and learning disability)(5,6) and neuronal environmental insults (chronic increased systemic inflammation and history of autoimmune disease)(7) were associated with early-onset and non-amnestic forms of dementia (primary progressive aphasias). In the present study, we investigated these same neurodevelopmental and neuroenvironmental factors in a large heterogenous AD cohort, to determine if they influence symptom age at onset and/or the phenotypic presentation of AD.

## Methods

### Discovery Cohort

Searching the University of California San Francisco (UCSF) Memory and Aging Center (MAC) database, we identified n=2652 subjects from 1998-2016 who met diagnostic (NINCDS-ADRDA and later NIA-AA) criteria for probable AD.(8,9) EOAD and LOAD status were determined based on the age at first symptom with EOAD defined as <65 and LOAD ≥65. Non-amnestic AD classifications were restricted to the diagnosis of logopenic variant primary progressive aphasia (lvPPA)(10) or posterior cortical atrophy (PCA),(11) given the degree of AD clinicopathological correlation. Individuals were excluded if they possessed clinical diagnoses of comorbid Lewy Body, Parkinson’s disease, and/or significant evidence of vascular disease defined by SIVD criteria(12) or if they possessed incomplete charts (defined as missing age at first symptoms, hand preference, past medical history, and/or social history). Also excluded were subjects in whom amyloid biomarker data or an autopsy pathology report was inconsistent with a diagnosis of underlying AD, or if an individual possessed a known autosomal dominant genetic cause for dementia, even if that gene was a known cause of AD, as the goal of this study was to investigate factors that impact idiopathic forms of disease. Lastly, we excluded cases whose age at onset was greater than age 85 as knowledge about AD epidemiology in populations older than 85 is limited and begins to demonstrate much greater amounts of co-pathology.(13,14) Splitting into EOAD and LOAD cohorts, we included 750 consecutive cases of EOAD and then case matched by number, LOAD cases (**Figure 1**).

**Figure 1.**
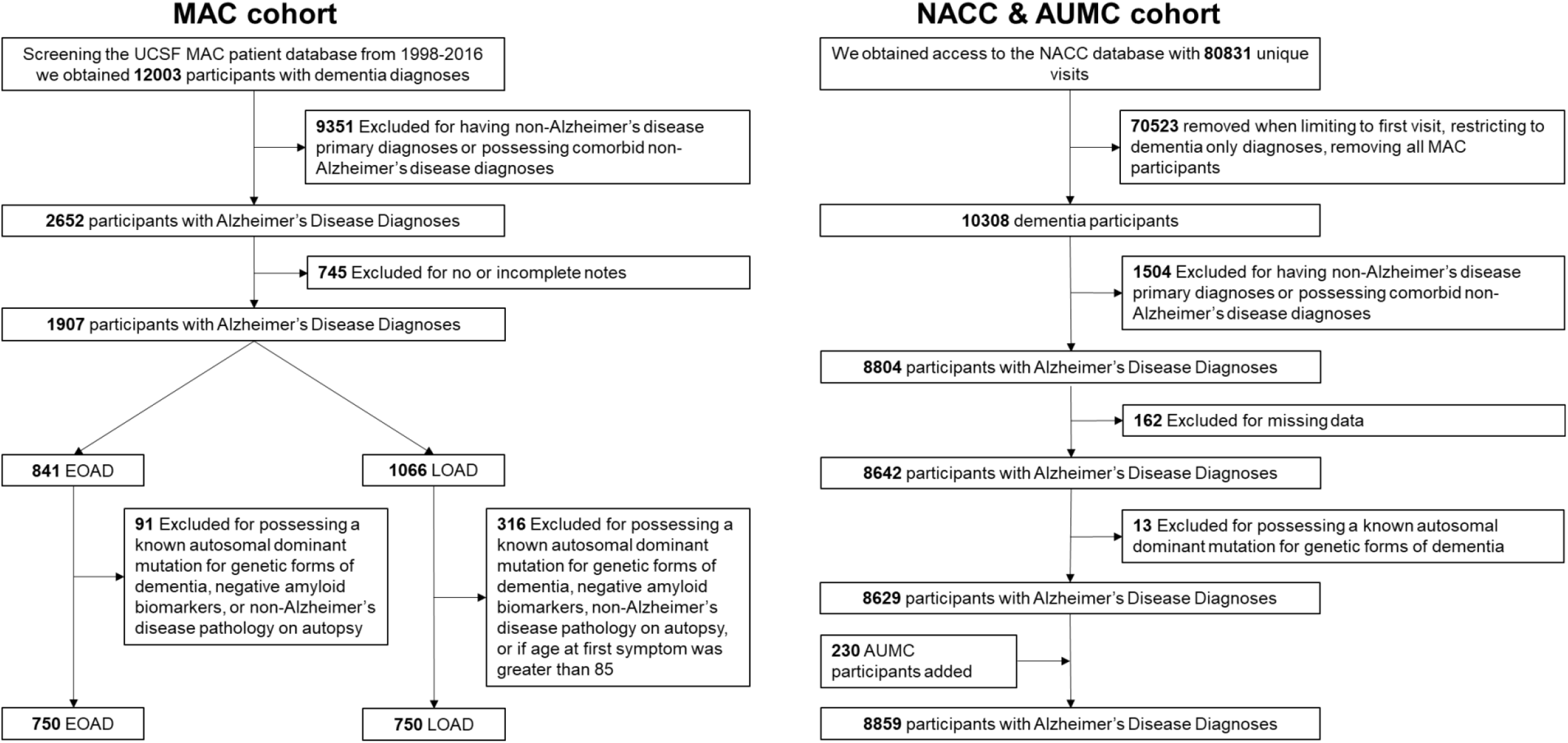
MAC and NACC & AUMC AD Cohort Flowchart. As our intention was to study individuals with sole predicted underlying AD pathology, subjects were excluded if they possessed 1) clinical diagnoses of comorbid Lewy Body, Parkinson’s disease, and/or significant evidence of vascular disease defined by SIVD criteria,(12) 2) incomplete charts (defined as missing age at first symptoms, hand preference, past medical history, and/or social history, 3) known autosomal dominant AD and/or FTD mutations (*APP, C9ORF72, FUS, GRN, MAPT, PSEN1*, and *PSEN2*), and or diagnoses of Down’s syndrome, 4) when available, were negative for AD biomarkers (CSF, Amyvid, and/or PiB PET) and/or AD pathology at autopsy,(69) or 5) were older than age 85 at first symptoms, as the epidemiology is understudied but believed to differ from typical amnestic AD, possessing a greater amount of multiple neurodegenerative disease co-pathologies.(13,14)

### External Validation Cohorts

Selecting individuals meeting criteria for dementia with an etiology of Alzheimer’s disease from the National Alzheimer’s Coordinating Center (NACC) database,(15,16) filtering out MAC cases, incomplete charts, and known autosomal dominant disease-causing mutations, produced 8,629 subjects from September 2005-June 2016. As the NACC database lacked sufficient numbers of non-amnestic AD we obtained 230 additional subjects from Amsterdam University Medical Center (AUMC) (36 lvPPA, 61 PCA, 66 EOAD, 67 LOAD), increasing the total to 8,859 (**Figure 1**). For comparison of autoimmune prevalence, specifically, we supplemented the AUMC group with cases from Mayo Clinic, Jacksonville (MCJ) (43 lvPPA, 81 PCA). The MCJ participants were not incorporated in the above 8,859 collection, to avoid the possibility of co-enrollment in the NACC.

### Standard Protocol Approvals, Registrations, and Patient Consents

Human research committees at the MAC, AUMC, and MCJ as well as all participating NACC sites approved the study of patients’ clinical data. Written informed consent from participants or responsible surrogates was obtained in accordance with the Declaration of Helsinki.

### Identification and Classification of Typical AD Risk and Novel Factors

At each site, charts were reviewed, screening for hypertension, hypercholesterolemia, diabetes, handedness, learning disability,(5) and seizure (as defined by the NACC: “active” if the event occurred within the past year or still required active management, “remote” if the event occurred over one year ago and completely resolved or is without ongoing management), and autoimmune disease(5,17) (**Supplemental Table 1**).

## Data Availability

All data used in this study are available for review upon formal request. As the institutional procedures in place at the time participants’ gave informed consent do not authorize open data sharing, all requests will need to undergo UCSF MAC regulated procedures including the submission of a materials transfer agreement. The requesting party will need to provide their name and affiliation as well as a brief description of their intended use of the data.

### Data Analyses

Data were summarized for EOAD and LOAD groups using means and standard deviations for continuous variables and counts and proportions for categorical data. For comparisons between EOAD and LOAD, analysis of variance or Students independent groups t-tests were used for continuous variables such as age, and education and Fisher’s exact or Chi-squared tests for nominal variables such as sex, non-right-handedness, learning disability, or presence of autoimmune disease. Odds ratios between EOAD and LOAD for risk factors were calculated using logistic regression controlled for significant covariates. For validation of the groupings of these risk factors within EOAD and LOAD, principal component analysis identified composite risk factors by EOAD and LOAD using the Kaiser criterion (principal component eigenvalues > 1.0) with loadings above 0.4 or below -0.4 as cutoffs used to identify variable importance in each identified factor. In addition, we created heatmaps using the Krakov visual techniques and odds ratios to identify differences and similarities between the cohorts.(18) The heatmaps were compared visually to identify similarities and differences and using chi-squared tests to test for independence. All analyses were performed with STATA 17.1 with significance levels set to *p*<0.05. Although the quantitative analyses involved many combinations of outcomes and predictors, we did not perform formal adjustments for multiple comparisons for each of the factors. This was because: a) the hypotheses were highly specific; and b) we expected many measures to show statistically significant differences between groups and the directions and magnitudes of the differences could fit a biologically coherent pattern with each result reinforcing the other, rather than detracting from one another, as required by formal multiple comparison adjustments such as Bonferroni.(19,20)

## Results

Within the MAC cohort, EOAD showed significantly higher proportions of non-right-handedness, learning disability, seizure, autoimmune disease, and non-amnestic AD presentations, while LOAD demonstrated higher frequencies of hypertension, hypercholesterolemia, and diabetes (*p*<0.001 for all except remote seizure, *p*=0.002, and autoimmune disease, *p*=0.007). Sex, education, and *APOE*-ɛ4 allelic frequencies were no different between groups. External cohorts, from the NACC & AUMC replicated increased non-right-handedness and seizure within EOAD and increased proportions of hypertension, hypercholesterolemia, and diabetes associated with LOAD (*p*<0.001 for all comparisons except handedness, *p*=0.02). NACC & AUMC LOAD showed a higher proportion of women, fewer years of education, and decreased *APOE*-ɛ4 allelic frequencies compared to EOAD (*p*<0.001 for all comparisons) (**Table 1**). To confirm these results we performed logistic regressions predicting EOAD vs. LOAD controlling for sex, education, *APOE*-ɛ4 status, typical, and novel risk factors, which confirmed all significant analyses (**Table 2**).

**Table 1.**
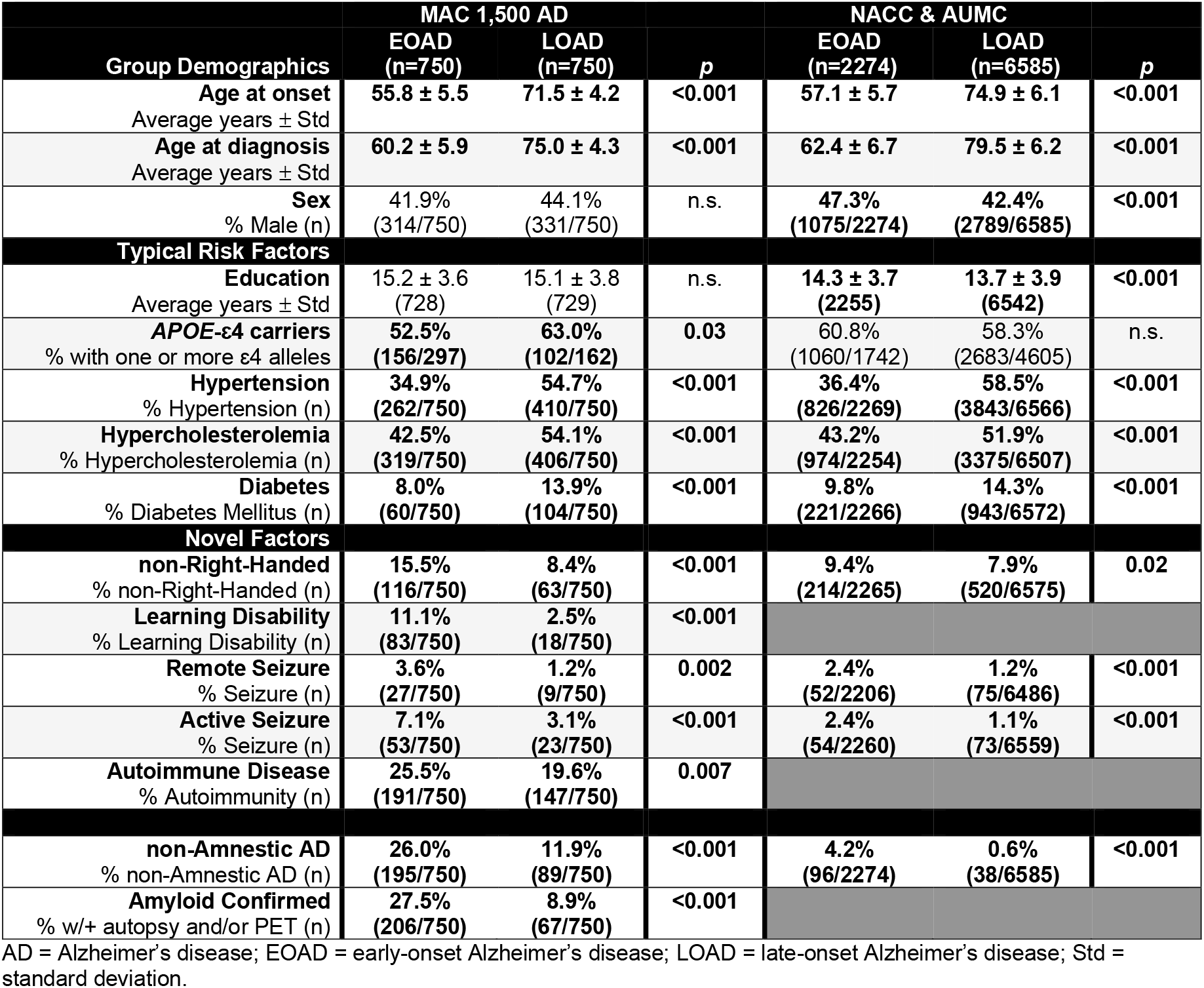
Demographics, Typical, and Novel Factors in MAC and NACC & AUMC in EOAD vs. LOAD.

**Table 2.** Logistic Regressions of Factors in MAC and NACC & AUMC that Predict EOAD vs. LOAD.

|  | MAC 1,500 AD<br>95% C.I. for OR |  |  |  | *p* | NACC & AUMC<br>95% C.I. for OR |  |  |  | *p* |
| --- | --- | --- | --- | --- | --- | --- | --- | --- | --- | --- |
|  | OR | Lower | Upper |  |  | OR | Lower | Upper |  |  |
| Education | 1.022 | 0.953 | 1.095 |  | n.s. | 0.882 | 0.774 | 1.004 |  | n.s. |
| *APOE*-ε4 carriers | 0.674 | 0.437 | 1.038 |  | n.s. | 1.109 | 0.508 | 2.422 |  | n.s. |
| **Decreased EOAD Prevalence** | **OR** | **Lower** | **Upper** |  | ***p*** | **OR** | **Lower** | **Upper** |  | ***p*** |
| Hypertension | 0.445 | 0.362 | 0.548 |  | <0.001 | 0.405 | 0.367 | 0.446 |  | <0.001 |
| Hypercholesterolemia | 0.627 | 0.511 | 0.836 |  | <0.001 | 0.706 | 0.641 | 0.777 |  | <0.001 |
| Diabetes Mellitus | 0.540 | 0.386 | 0.755 |  | <0.001 | 0.643 | 0.551 | 0.751 |  | <0.001 |
| **Increased EOAD Prevalence** | **OR** | **Lower** | **Upper** |  | ***p*** | **OR** | **Lower** | **Upper** |  | ***p*** |
| non-Right-Handed | 2.029 | 1.464 | 2.816 |  | <0.001 | 1.218 | 1.031 | 1.439 |  | 0.020 |
| Learning Disability | 5.051 | 3.012 | 8.547 |  | <0.001 |  |  |  |  |  |
| Remote Seizure | 2.677 | 1.150 | 5.759 |  | 0.012 | 2.183 | 1.506 | 3.163 |  | <0.001 |
| Active Seizure | 1.953 | 1.150 | 3.315 |  | 0.013 | 2.315 | 1.506 | 3.163 |  | <0.001 |
| Autoimmune Disease | 1.587 | 1.064 | 2.364 |  | 0.023 |  |  |  |  |  |

AD = Alzheimer's disease; C.I. = confidence interval; EOAD = early-onset Alzheimer's disease; LOAD = late-onset Alzheimer's disease; OR = odds ratio; Std = standard deviation.

To determine if these results were mediated by the increased proportion of non-amnestic AD within EOAD, we separated out the non-amnestic AD cases from the total 1,500 AD cohort, and all prior EOAD and LOAD differences survived. Comparing the non-amnestic AD cohort against the remaining amnestic EOAD and amnestic LOAD revealed statistically lower *APOE*-ɛ4 allelic frequencies in non-amnestic AD (*p*<0.001) (**Supplemental Table 2**). As a further sub-analysis, we split the non-amnestic AD cohort into its constituent groups, lvPPA and PCA. The PCA cohort was significantly younger at age at onset and age of diagnosis, possessed a lower frequency of diabetes, and higher rates of active seizure, and autoimmune disease (**Supplemental Table 3**).

As these groups were based on the clinical diagnosis of an AD syndrome, we performed analyses within each of the diagnoses (non-amnestic AD, amnestic EOAD, amnestic LOAD) broken down by cases with and without confirmed amyloid disease (autopsy proven and/or amyloid PET biomarker positivity) to determine how well our results generalized to pathologically proven AD. Non-amnestic AD and amnestic EOAD amyloid confirmed cases were younger at age of onset and age of first visit than their unconfirmed counterparts. Consistent with this younger age, they also possessed relative decreases in vascular risk factors. There was a greater proportion of male participants in amyloid confirmed amnestic EOAD and LOAD groups than those without amyloid confirmation and across all diagnoses, years of education was higher in the amyloid confirmed group. There were no statistically significant differences among the novel factors between amyloid confirmed and unconfirmed groups, except for an even greater amount of learning disability within the amyloid confirmed non-amnestic AD (**Supplemental Table 4**).

Reducing the novel and typical risk factors into single composites (positive for typical factors if hypertension, hypercholesterolemia, and/or diabetes were present, and for novel factors only if non-right-handedness and/or seizure were present – as autoimmune disease and learning disability were not adequately collected in the NAAC, these categories were not considered in this analysis), within a combined MAC/NACC & AUMC cohort, histogram plots revealed a mean age at onset of 66.9 years for novel factors, 71.4 years for typical AD risk factors, and 69.2 as the average difference between these two (**Figure 2**).

**Figure 2.**
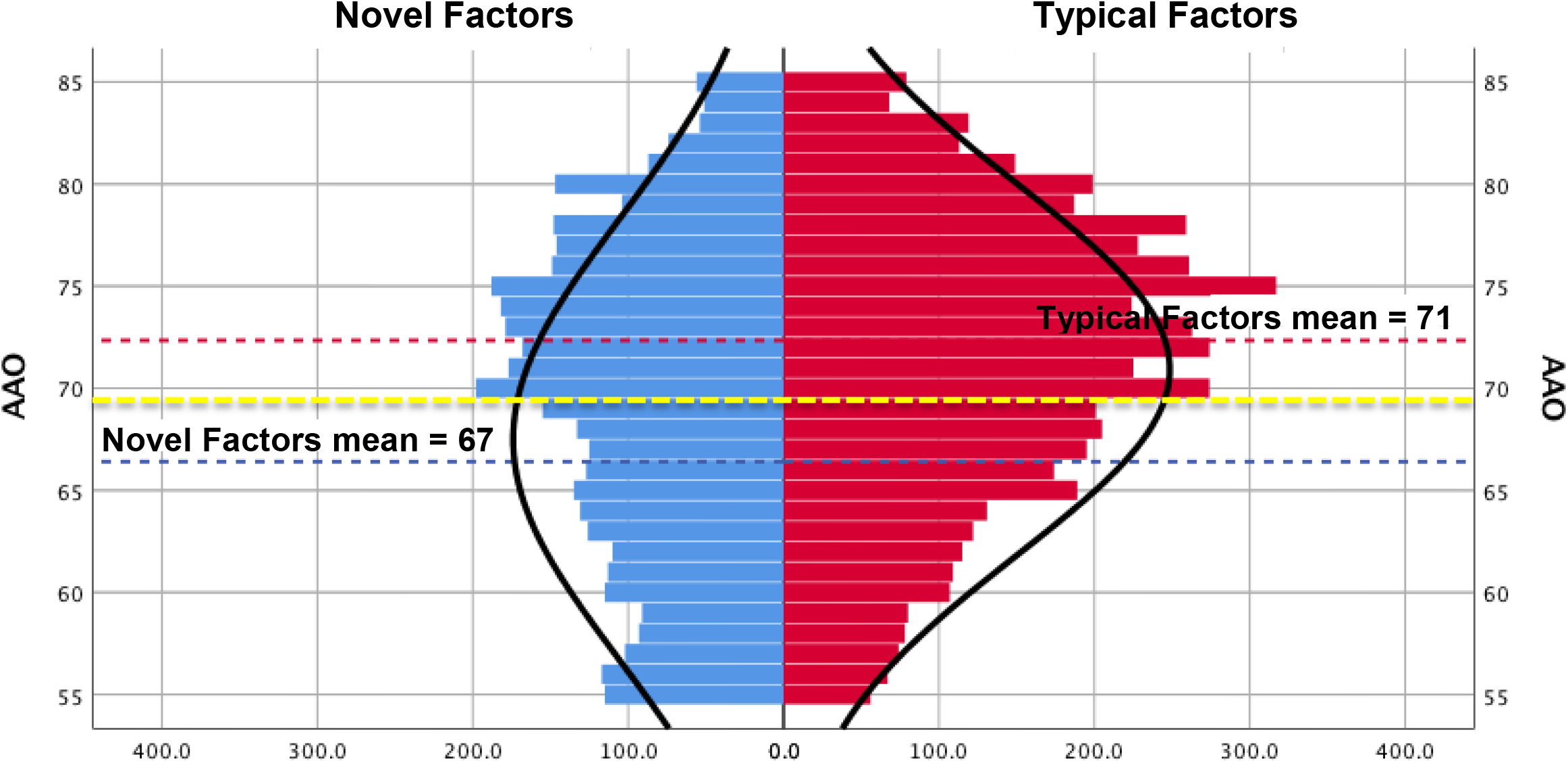
Histograms of AD Age at Onset by Novel vs. Typical Factors. Plotting the MAC, NACC, and AUMC individuals who possessed novel factors (only non-right-handedness and seizure were assessed, as these were the only novel factors consistently collected across the various cohorts) in blue, and typical AD risk factors (hypertension, hypercholesterolemia, and diabetes mellitus) on the right, in red, by 2 year AAO epochs, we obtained distribution curves for each. The mean AAO for novel factors was 66.9 years (blue dashed line), for typical factors, 71.4 years (red dashed line), and these distributions differed statistically (*p*=0.003). Taking the mean of these two means, 69.2 years (yellow dashed line), maximally distinguishes the amounts of novel factors vs. typical AD risk factors within this cohort.

To determine how novel factors interacted with the most significant AD risk factor, *APOE*-ɛ4, we isolated the subset of individuals who were positive for a novel factor and had undergone *APOE*-ɛ4 testing, stratifying by *APOE*-ɛ4 positive and negative status. Further, as rates of *APOE*-ɛ4 differed in non-amnestic AD cohorts from the remaining EOAD and LOAD, we plotted the non-amnestic AD cohort separately. Here we found that within the non-amnestic AD cohort there were no age at onset differences between *APOE*-ɛ4 positive carriers and negative carriers, whereas in the remaining EOAD and LOAD, the interaction of *APOE*-ɛ4 with novel factors was significant (*p*=0.013), with *APOE*-ɛ4 positive carriers showing first symptoms on average 3 years before than those who were *APOE*-ɛ4 negative (**Supplemental Figure 1**).

As age at onset determination is inherently imprecise (based on clinician report of the patient’s or informant’s testimony), we split each cohort into quintiles to investigate how various factors displayed across age at onset. With this, novel factors followed inverse linear relationships with age at onset, while the typical factors mostly fit quadratic trends (**Figure 3, Supplemental Table 5**). To determine the degree of burden of each factor, we divided the MAC cohort by individual factor of interest and found that non-right-handedness, learning disability, active seizure, remote seizure, and autoimmune groups, were all significantly younger at age at onset (*p*<0.001, except autoimmune disease, *p*<0.05) (**Supplemental Table 6**). Principal component analysis loaded all factors into three components, one consisting of the typical AD risk factors (hypertension, hypercholesterolemia, and diabetes) and two separating out novel factors non-right-handedness and learning disability from autoimmune disease and seizure. As offered above, within the NACC collection, among the novel factors, only non-right-handedness and seizure history, were adequately captured. As non-right-handedness and seizure reflected factors within each of the two components identified in the MAC cohort, we combined the MAC and NACC & AUMC cohorts, restricting analyses to non-right-handedness and seizures. Survival curve analysis demonstrated that the age at which 50% of those with only one novel factor (non-right-handedness or seizure) developed first AD symptoms was 3 years younger than those without either factor (68 vs. 71). The group with both novel factors (non-right-handedness and seizure) was nine years younger than those with only one single novel factor (59 vs. 68) and twelve years younger than those with neither (59 vs. 71) (log-rank *p*<0.001 across all comparisons using ANOVA with a Scheffe correction for multiple comparisons) (**Figure 4 and Supplemental Figure 2**).

**Figure 3.**
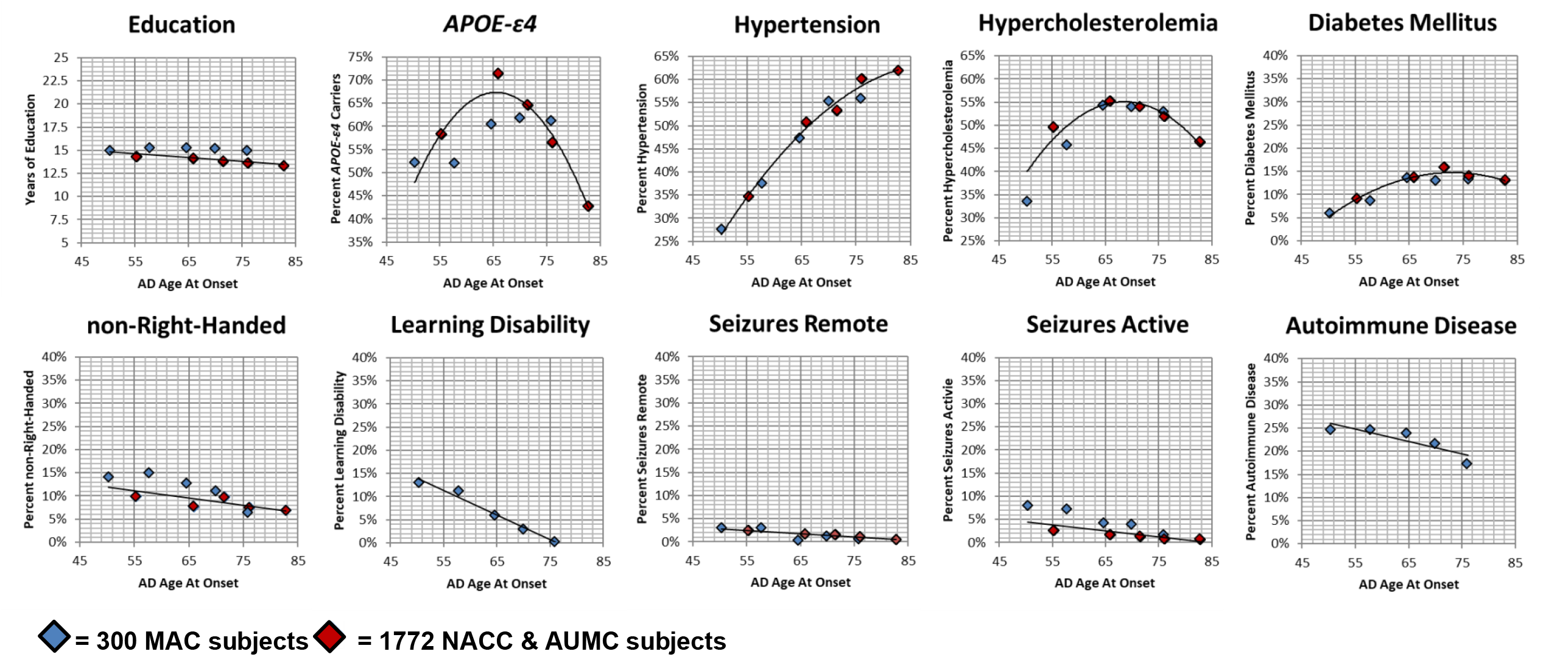
MAC and NACC & AUMC AD Distribution of Factors Across Age at Onset Split into Quintiles. Each diamond represents a quintile with its respective cohort, with each blue diamond reflecting the 300 MAC subjects and each red diamond, the NACC & AUMC cohort. Each diamond is plotted out with the average of that specific cohort by the percent of a specific factor, like non-right-handedness. The exact numerical values for each quintile can be found in supplemental material (**Supplemental Table 3**). Accounting for weighted differences between the red and blue diamonds, best-fit lines were generated. The top row consists of factors already known to impact AD risk, typical AD risk factors, while the bottom row reflects the series novel factors we investigated in this study.

**Figure 4.**
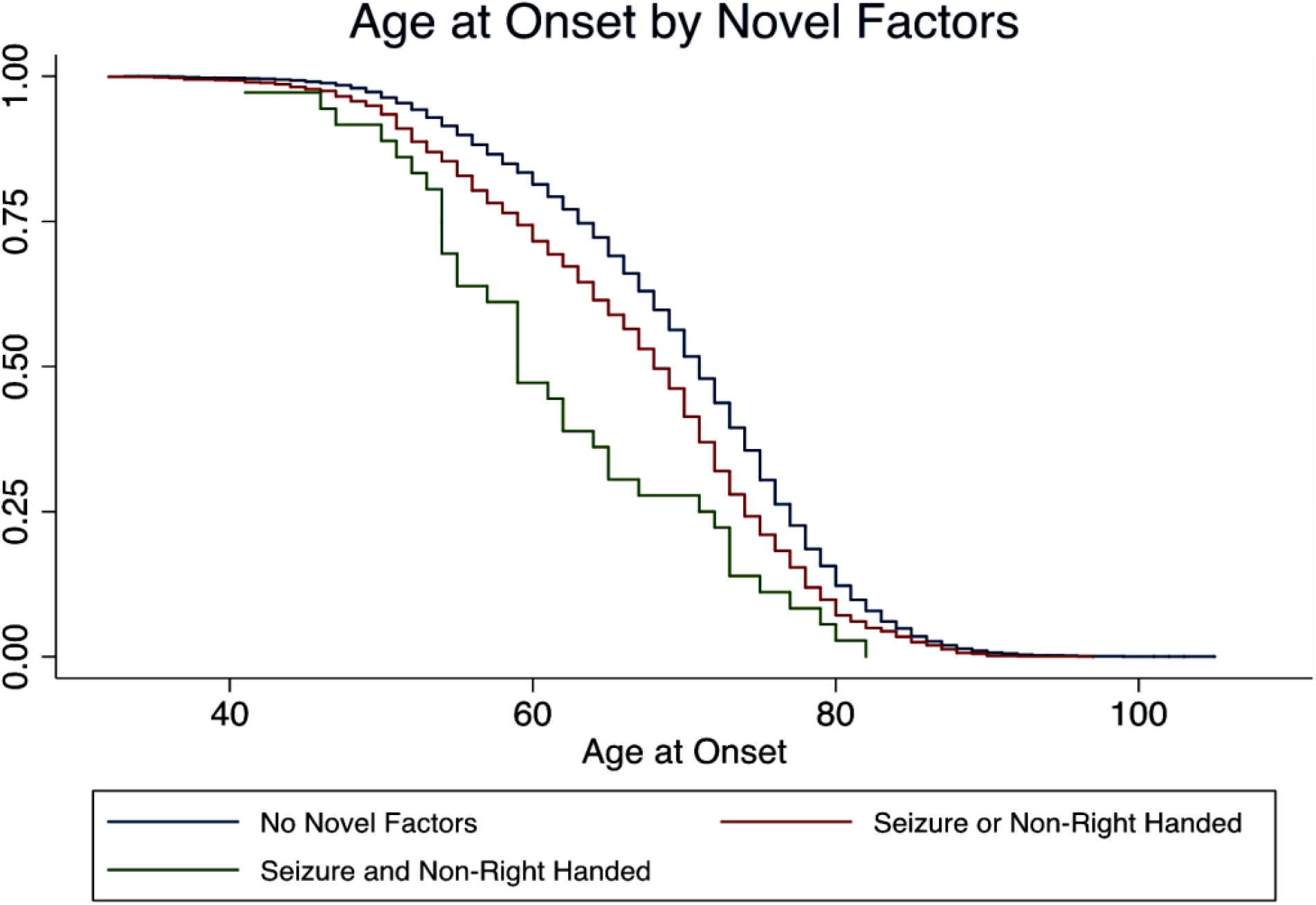
Burden of Factors Seizure and non-Right-Handedness on AD Age at Onset. In combined MAC and NACC & AUMC cohorts, plotting individuals stratified by numbers of novel factors (non-right-handedness and seizure) versus the age at which they developed first symptoms produced three distinct Kaplan-Meier curves. The blue line (n=9074) encompasses all participants who lacked any novel factor. The red line (n=1190) included only those who had one novel factor (either non-right-handedness or seizure). The green line (n=36) comprised those individuals who possessed both novel factors (non-right-handedness and seizure). The age at which 50% of individuals with no novel factors develops first symptoms is 71, with only one factor is 68, and with both factors is 59. Using ANOVA with a Scheffe correction for multiple comparisons: No Novel factors vs. one Novel factor (blue vs. red) *p*<0.001; No Novel factors vs. two Novel factors (blue vs. green) log rank *p*<0.001; One Novel factor vs. two Novel factors (red vs. green) *p*=0.009.

To display the relative contributions of autoimmunity within the various AD cohorts, we created heat maps reflecting the prevalence of individual autoimmune conditions compared to general population values. Autoimmune conditions displayed a gradient of prevalence where both the number of distinct autoimmune disorders as well as the degree of their overrepresentation was greatest in non-amnestic AD, followed by EOAD, and least in LOAD. Within the MAC cohort the top six overrepresented autoimmune conditions (greater than five times general population estimates) were also identified within the AUMC & MCJ cohort, at near identical elevated rates (**Table 3**). In support of this qualitative assessment, formal chi-squared analysis of the top six conditions in the MAC cohort and within the AUMC & MCJ cohort was not significant, indicating they are not independent (*p*=0.544).

**Table 3.** Autoimmune Prevalence & Estimated Odds Ratios in MAC and AUMC & MCJ Across Total AD Cohort and Subtypes.

| Autoimmune disease | MAC 1,500 AD Cohort |  |  |  | General Population Prevalence | AUMC & MCJ |  |  |  |
| --- | --- | --- | --- | --- | --- | --- | --- | --- | --- |
|  | Total (n=1500) | non-Amnesic (n=284) | EOAD (n=555) | LOAD (n=661) |  | Total (n=354) | non-Amnesic (n=221) | EOAD (n=66) | LOAD (n=67) |
| **ANCA associated vasculitis**<br>(prevalence)<br>(n) | **23**<br>0.07%<br>1 | n/a<br>0.00%<br>0 | **60**<br>0.18%<br>1 | n/a<br>n/a<br>0 | 0.003%(54) | **93**<br>0.28%<br>1 | n/a<br>0.00%<br>0 | **505**<br>1.50%<br>1 | n/a<br>n/a<br>0 |
| **Guillain-Barré Syndrome**<br>(prevalence)<br>(n) | **17**<br>0.33%<br>5 | **53**<br>1.06%<br>3 | **18**<br>0.36%<br>2 | n/a<br>n/a<br>0 | 0.02%(55) | **14**<br>0.28%<br>1 | **23**<br>0.45%<br>1 | n/a<br>n/a<br>0 | n/a<br>n/a<br>0 |
| **Systemic Lupus Erythematosus**<br>(prevalence)<br>(n) | **14**<br>0.27%<br>4 | **18**<br>0.35%<br>1 | n/a<br>n/a<br>0 | **23**<br>0.45%<br>3 | 0.02%(56) | **28**<br>0.56%<br>2 | **45**<br>0.90%<br>2 | n/a<br>n/a<br>0 | n/a<br>n/a<br>0 |
| **Idiopathic Thrombocytopenic Purpura**<br>(prevalence)<br>(n) | **13**<br>0.13%<br>2 | **35**<br>0.35%<br>1 | **18**<br>0.18%<br>1 | n/a<br>n/a<br>0 | 0.01%(57) | **28**<br>0.28%<br>1 | **45**<br>0.45%<br>1 | n/a<br>n/a<br>0 | n/a<br>n/a<br>0 |
| **Multiple Sclerosis**<br>(prevalence)<br>(n) | **9**<br>0.53%<br>8 | **29**<br>1.76%<br>5 | **6**<br>0.36%<br>2 | **3**<br>0.15%<br>1 | 0.06%(56) | **9**<br>0.56%<br>2 | **15**<br>0.90%<br>2 | n/a<br>n/a<br>0 | n/a<br>n/a<br>0 |
| **Sarcoidosis**<br>(prevalence)<br>(n) | **7**<br>0.27%<br>4 | **9**<br>0.35%<br>1 | **9**<br>0.36%<br>2 | **4**<br>0.15%<br>1 | 0.04%(58) | **7**<br>0.28%<br>1 | **11**<br>0.45%<br>1 | n/a<br>n/a<br>0 | n/a<br>n/a<br>0 |
| **Systemic Sclerosis/Scleroderma**<br>(prevalence)<br>(n) | **6**<br>0.13%<br>2 | **15**<br>0.35%<br>1 | **8**<br>0.18%<br>1 | n/a<br>n/a<br>0 | 0.02%(59) | n/a<br>n/a<br>0 | n/a<br>n/a<br>0 | n/a<br>n/a<br>0 | n/a<br>n/a<br>0 |
| **Myasthenia Gravis**<br>(prevalence)<br>(n) | **3**<br>0.07%<br>1 | **18**<br>0.35%<br>1 | n/a<br>n/a<br>0 | n/a<br>n/a<br>0 | 0.02%(60) | n/a<br>n/a<br>0 | n/a<br>n/a<br>0 | n/a<br>n/a<br>0 | n/a<br>n/a<br>0 |
| **Crohn's Disease**<br>(prevalence)<br>(n) | **3**<br>0.60%<br>9 | **5**<br>1.06%<br>3 | **2**<br>0.36%<br>2 | **3**<br>0.61%<br>4 | 0.20%(61) | n/a<br>n/a<br>0 | n/a<br>n/a<br>0 | n/a<br>n/a<br>0 | n/a<br>n/a<br>0 |
| **Ulcerative Colitis**<br>(prevalence)<br>(n) | **2**<br>0.53%<br>8 | **4**<br>1.06%<br>3 | n/a<br>n/a<br>0 | **3**<br>0.76%<br>5 | 0.24%(61) | **4**<br>0.85%<br>3 | **4**<br>0.90%<br>2 | **6**<br>1.50%<br>1 | n/a<br>n/a<br>0 |
| **Pernicious Anemia**<br>(prevalence)<br>(n) | **2**<br>0.27%<br>4 | **2**<br>0.35%<br>1 | **1**<br>0.18%<br>1 | **2**<br>0.30%<br>2 | 0.15%(56) | **4**<br>0.56%<br>2 | **6**<br>0.90%<br>2 | n/a<br>n/a<br>0 | n/a<br>n/a<br>0 |
| **Type I Diabetes Mellitus**<br>(prevalence)<br>(n) | **2**<br>0.40%<br>6 | n/a<br>0.00%<br>0 | **5**<br>0.90%<br>5 | **1**<br>0.15%<br>1 | 0.19%(56) | **2**<br>0.28%<br>1 | **2**<br>0.45%<br>1 | n/a<br>n/a<br>0 | n/a<br>n/a<br>0 |
| **Hypothyroidism**<br>(prevalence)<br>(n) | **2**<br>15.27%<br>229 | **2**<br>16.90%<br>48 | **2**<br>16.40%<br>91 | **2**<br>13.62%<br>90 | 6.90%(62) | **1**<br>7.34%<br>26 | **1**<br>9.50%<br>21 | **0.4**<br>3.00%<br>2 | **0.6**<br>4.48%<br>3 |
| **Sjögren's Syndrome**<br>(prevalence)<br>(n) | **1**<br>0.47%<br>7 | **1**<br>0.35%<br>1 | **2**<br>0.54%<br>3 | **1**<br>0.45%<br>3 | 0.32%(56) | **n/a**<br>n/a<br>0 | **n/a**<br>n/a<br>0 | **n/a**<br>n/a<br>0 | **n/a**<br>n/a<br>0 |
| **Celiac Disease**<br>(prevalence)<br>(n) | **1**<br>0.47%<br>7 | **n/a**<br>0.00%<br>0 | **1**<br>0.54%<br>3 | **1**<br>0.60%<br>4 | 0.83%(63) | **n/a**<br>n/a<br>0 | **n/a**<br>n/a<br>0 | **n/a**<br>n/a<br>0 | **n/a**<br>n/a<br>0 |
| **Ankylosing Spondylitis/HLA B27**<br>(prevalence)<br>(n) | **1**<br>0.33%<br>5 | **1**<br>0.35%<br>1 | **0.4**<br>0.18%<br>1 | **1**<br>0.45%<br>3 | 0.50%(59) | **n/a**<br>n/a<br>0 | **n/a**<br>n/a<br>0 | **n/a**<br>n/a<br>0 | **n/a**<br>n/a<br>0 |
| **Graves Thyroiditis**<br>(prevalence)<br>(n) | **1**<br>1.60%<br>24 | **1**<br>1.76%<br>5 | **1**<br>1.80%<br>10 | **1**<br>1.36%<br>10 | 2%(62) | **n/a**<br>n/a<br>0 | **n/a**<br>n/a<br>0 | **n/a**<br>n/a<br>0 | **n/a**<br>n/a<br>0 |
| **Rheumatoid Arthritis**<br>(prevalence)<br>(n) | **1**<br>1.27%<br>19 | **1**<br>0.70%<br>2 | **2**<br>1.44%<br>8 | **2**<br>1.36%<br>9 | 0.86%(56) | **1**<br>0.85%<br>3 | **0.5**<br>0.45%<br>1 | **4**<br>3.00%<br>2 | **n/a**<br>n/a<br>0 |
| **Psoriasis**<br>(prevalence)<br>(n) | **0.8**<br>1.13%<br>17 | **1.2**<br>1.76%<br>5 | **1.1**<br>1.62%<br>9 | **0.4**<br>0.60%<br>4 | 1.5%(64) | **1.5**<br>2.26%<br>8 | **2.1**<br>3.17%<br>7 | **1.0**<br>1.50%<br>1 | **n/a**<br>n/a<br>0 |
| **Discoid Lupus**<br>(prevalence)<br>(n) | **0.3**<br>0.27%<br>4 | **n/a**<br>n/a<br>0 | **0.2**<br>0.18%<br>1 | **1**<br>0.45%<br>3 | 0.80%(65) | **n/a**<br>n/a<br>0 | **n/a**<br>n/a<br>0 | **n/a**<br>n/a<br>0 | **n/a**<br>n/a<br>0 |
| **Vitiligo**<br>(prevalence)<br>(n) | **0.3**<br>0.13%<br>2 | **n/a**<br>n/a<br>0 | **1**<br>0.36%<br>2 | **n/a**<br>n/a<br>0 | 0.40%(56) | **0.7**<br>0.28%<br>1 | **1**<br>0.45%<br>1 | **n/a**<br>n/a<br>0 | **n/a**<br>n/a<br>0 |
| **Lichen Sclerosus**<br>(prevalence)<br>(n) | **0.2**<br>0.07%<br>1 | **1**<br>0.35%<br>1 | **n/a**<br>n/a<br>0 | **n/a**<br>n/a<br>0 | 0.30%(66) | **1**<br>0.28%<br>1 | **2**<br>0.45%<br>1 | **n/a**<br>n/a<br>0 | **n/a**<br>n/a<br>0 |
| **Lupoid Hepatitis**<br>(prevalence)<br>(n) | **n/a**<br>n/a<br>0 | **n/a**<br>n/a<br>0 | **n/a**<br>n/a<br>0 | **n/a**<br>n/a<br>0 | 0.02%(67) | **14**<br>0.28%<br>1 | **23**<br>0.45%<br>1 | **n/a**<br>n/a<br>0 | **n/a**<br>n/a<br>0 |
| **Polymyalgia Rheumatica\***<br>(prevalence)<br>(n) | **n/a**<br>n/a<br>0 | **n/a**<br>n/a<br>0 | **n/a**<br>n/a<br>0 | **n/a**<br>n/a<br>0 | 1.00%(68) | **1**<br>1.13%<br>4 | **0.5**<br>0.45%<br>1 | **n/a**<br>n/a<br>0 | **4**<br>4.48%<br>3 |

Each colored cell displays the estimated odds ratio, in bold, on top, its prevalence, in the middle, and numerical count on the bottom. Autoimmune diseases were rank ordered by the estimated odds ratio within the total MAC cohort. Disorders not observed in the MAC cohort, were similarly ordered in the AUMC & MCJ. The color coding within each cell reflects the estimated odds ratio for that particular autoimmune condition: Red $\geq 20x$ or greater; tangerine/dark orange = 10-19x; light orange = 5-9x; yellow = 3-4x; green = 1-2x; blue < 1x; grey = none observed/not applicable. Both datasets revealed similar patterns of autoimmunity within total AD cohorts as well as across subcohorts.

## Discussion

In this study, we explored the demographic and health histories of early onset and non-amnestic AD compared to late onset AD, as a means of uncovering novel factors enriched in these rarer forms of AD. We discovered that non-right-handedness, learning disability, seizure, and autoimmune disease each influenced the age at onset and phenotypical targeting of AD. We proceeded to validate these findings by employing external cohorts from the NACC, AUMC, and MCJ, that increased our total investigated cohort to over 10,000 AD participants. While past reports have demonstrated aspects of the relationship between AD and some of the factors we studied,(5,21–23) this report is the largest and most comprehensive analysis of its kind. Additionally, the mounting contemporary evidence ascribing biological relevance to each of these factors in the pathophysiology of AD warranted their modern in-depth investigation. For example, until recently the genetics of non-right-handedness remained elusive but now have been shown to possess strong association with risk of neurodegenerative disease, including AD.(24) Our own work demonstrated a domain-specific association between developmental differences and phenotypical presentation of AD(6) as well as identified localized cortical dysplastic changes in lvPPA with a history of developmental dyslexia in a follow up autopsy study, suggesting that focal neuronal migratory irregularities might link together the presence of early life learning disability with later life phenotypical targeting of neurodegenerative disease.(25) Studies of adults with a history of childhood-onset epilepsy observed increased β-amyloid burden in midlife compared to controls, demonstrating the potential contribution seizure activity has towards AD pathophysiology.(26) Similarly, midlife chronic inflammation has been shown to predispose individuals to later development of dementia.(27) Finally, individuals who suffered from particular autoimmune diseases presented with an increased risk of AD, that was subsequently diminished if the autoimmune disease was treated with immunotherapy.(28) Taken together, the elevated rates of non-right-handedness, learning disability, seizure, and autoimmune disease we observed within early onset and non-amnestic AD support a new theoretical framework for capturing individualized neurodegenerative disease susceptibility risk, which we discuss in detail below.

While we began this study by exploring the relationships of novel and typical AD risk factors across the EOAD age cutoff of <65, given the rather capricious origins of the EOAD age divide,(2) we quickly appreciated that our study design provided the additional opportunity to test the merits of this cutoff itself and model an age divide that optimized differences between these risk factors. Instead of finding support for the current age cutoff of age <65, we found that an age at first symptom of <70 maximally distributed all factors, established and novel, across the cohort (**Figure 2**). A prior study investigating differences in AD neuropsychological performances, came to similar conclusions, that a cutoff of <70 better distinguished the cognitive profiles observed within EOAD and LOAD than <65.(29) As AD prevalence doubles every 5 years after age 65,(3) if a new definition of <70 were to be put in place of the current <65 cutoff, it would triple the world’s population of EOAD, turning a rare disorder into one that comprises nearly 1/3 of all AD presentations. Moreover, as our data argues that the disease mechanisms behind EOAD risk may differ from LOAD, with anticipated increases in access to early-life education along with decreases in cardiovascular disease burden, we speculate that the divide between EOAD and LOAD might actually be mutable, requiring periodic reevaluation. To this end, beyond a simplistic dichotomous divide, we also evaluated how each factor displayed across a range of ages. We observed inverse linear relationships across all novel factors and age at onset, such that differences presented between any two age ranges, even within the respective designations of EOAD and LOAD, adding even greater support for the relevance of these factors in modulating age at onset of AD. In contrast, typical AD risk factors better fit quadratic functions, aligning with prior studies that suggested typical AD risk factors were less relevant to the oldest AD presentations(30) (**Figure 3**). Combined, the distribution of these novel and typical factors across age ranges suggests that the current standards for AD age-based cutoffs deserve revisiting. Rather than maintaining a single EOAD/LOAD age divide, our data would suggest that it is perhaps more appropriate to classify AD individuals into one of three categories: early-onset (<70), typical-onset (70-85), and late-onset AD (>85).

Unlike prior studies on AD risk factors, typical and novel, here we not only investigated the prevalence of each individual factor, as detailed above, but also studied how they associated with each other.(31) Principle component analysis reduced non-right-handedness and learning disability into one factor and seizure and autoimmune disease into another. These relationships have substantial precedent, as increased prevalence of non-right-handedness with learning disability is well-described(31) and systemic chronic inflammation is a known instigator of seizure activity.(32) Further, the combination of these putative neurodevelopmental and neuroenvironmental factors produced even greater reductions in age at onset than predicted for each alone (**Figure 4 and Supplemental Figure 2**). The exponential decline in AD age at onset when both factors are present aligns with our prior work, which observed that neurodevelopmental differences(5,6) and chronic neuronal environmental insults(7) held unique status with regards to neurodegenerative disease susceptibility and supports the conceptualization of a ‘two-hit’ model of neurodegenerative disease where structurally, developmentally vulnerable regions become susceptible over time to chronic low-level autoinflammatory signals that in turn promote focal hyperexcitability, facilitating AD pathophysiology.

As to the exact inflammatory pathways mediating AD susceptibility, beyond the simple conceptualization of the presence or absence of autoimmune disease, we captured the specific autoimmune diagnoses when present and compared their prevalence rates in our cohort against general population estimates. Viewed in this manner, of the seven most overrepresented autoimmune conditions within the MAC AD cohort, the top six replicated in our external AUMC & MCJ cohort (**Table 3**). Moreover, three of these autoimmune disorders (systemic lupus erythematosus, idiopathic thrombocytopenic purpura, and multiple sclerosis) had previously been identified in association with dementia (including AD) in a large epidemiological study across the UK.(33) Mechanistically, all six of these overrepresented autoimmune disorders that presented in the MAC cohort and validated in our external AUMC & MCJ cohort share common etiologic associations with Fcγ receptor polymorphisms.(34,35) Fcγ receptors bridge adaptive and innate immune systems and show increased expression in microglia surrounding senile plaques.(36) Fcγ receptor-mediated phagocytosis pathways are implicated in the pathogenesis of AD(37) and abolishing Fcγ activity attenuates disease in AD mouse models.(38) The AD risk gene, *TREM2*, acts on microglia, triggering β-amyloid phagocytosis, through overlapping signaling with Fcγ receptors.(39) Thus, we hypothesize that the presence of certain autoimmune diseases might in part recapitulate *TREM2* mutation pathophysiology triggering an autoinflammatory signal that confers increased risk of AD. Moreover, pathological evidence shows that non-amnestic forms of AD display a higher degree of inflammatory microglial-associated changes than amnestic AD,(40) which is consistent with the gradient of autoimmune disease we observed across the MAC and AUMC & MCJ cohorts where non-amnestic disease displays both the greatest number and the greatest degree of overrepresentation of those conditions (**Table 3**).

In keeping with past reports, non-amnestic forms of AD were more frequent within our EOAD cohort.(3) Thus, in order to determine if the above associations between these novel and typical factors and AD age at onset were driven solely by the contribution of non-amnestic AD within our EOAD cohort, we compared EOAD and LOAD participants with non-amnestic AD phenotypes removed and proceeded to show that all prior differences survived (**Supplemental Table 2**). In the process of separating out non-amnestic AD presentations from amnestic EOAD and LOAD groups, we also confirmed past reports that rates of *APOE*-ɛ4 in non-amnestic, cortical-based presentations of AD were no different from the general population, but were elevated equally in amnestic, hippocampal-based AD, regardless of EOAD or LOAD status.(3) As novel factors were increased in both EOAD and non-amnestic AD groups, we further explored the effect that *APOE*-ɛ4 status in combination with the presence of novel factors had on age at onset. Here we found a dissociation, that in those who possessed a novel factor, *APOE*-ɛ4 positivity presented no additional effect on modulating age at onset in non-amnestic AD but had a significant effect on the remaining amnestic cases (**Supplemental Figure 1**). Thus, depending on AD disease phenotype, novel factors demonstrated effects that were independent of, or synergistic with, *APOE*-ɛ4 mechanisms of disease highlighting their relevance to AD risk modeling across age ranges and phenotypical presentations.

Separating out the non-amnestic forms of AD from the larger cohort also had the unintended consequence of highlighting the exceptional degree of novel risk factors present within this subgroup. PCA, in particular, showed the highest elevations of all novel factors (**Supplemental Table 2 and 3**). In this regard, it is of particular interest that a gene identified specifically as a PCA risk locus, *SEMA3C*,(41) is a member of a family of genes that have actions first in neurodevelopment and later in immune function.(42) Indeed, there is a growing literature that suggests prior to functioning within the immune system many notable immune-related genes act initially in shaping the developing brain, beginning with the observations that MHC Class I proteins first play roles in visual system radial glial migration (which is perhaps particularly relevant given that PCA is a disorder of visuoperceptive abilities).(43) Neurodevelopmental functions have now been described for Fcγ receptors(44) and *TREM2*.(45) Meanwhile, both neurodevelopmental differences and immune alterations have been appreciated with *APOE*-ɛ4.(46,47) Accordingly, it has been proposed that polymorphisms in genes that function in neurodevelopment and later act in maintaining cellular homeostasis, could produce conditions where developmental differences and later-life susceptibility to neurodegenerative disease co-occur, resulting in an age-dependent selective, focal attack on developmentally vulnerable brain regions.(25,48,49) A mechanism like this would most parsimoniously account for the exponential decrease in age at onset witnessed in the group of individuals who had both “hits” compared to those with only one “hit” (**Figure 4 and Supplemental Figure 2**), producing a nuance to the “two-hit” neurodegenerative hypothesis we put forward above, that in some discrete instances, individuals might possess a polymorphism in one of these particular genes that effectively encompasses a “two-in-one” hit. Further, this “two-in-one” hit hypothesis need not be restricted to neurodevelopmental and immune actions. In fact, due to the limitations of novel factor collections within our external validation cohort, we witnessed this exponential decrease in AD age at onset specifically in individuals who were both non-right-handed and suffered from seizure. This particular presentation aligns with the theories of pathological left-handedness, that in some instances non-right-handedness might come about as a consequence of early life brain injury. Most commonly observed in individuals suffering from epilepsy, pathological left-handedness also has theoretical association with learning disability among other conditions.(50) One study of a group of adults with epilepsy found an increased proportion of left-handedness in association with left and right parietal lobe injury.(51) Given that the left and right parietal lobes serve as the disease epicenters for lvPPA and PCA, respectively, our data highlights that there may be readily identifiable populations of individuals vulnerable to early-onset and non-amnestic Alzheimer’s disease, where the risks of developing focal cortical, atypical forms of AD could be predicted 4-5 decades beforehand, engendering potent opportunities for disease prevention.

Despite the scope of our undertaking and the extent to which our findings validated in external cohorts, this study has several limitations, most notably the data analyzed was largely derived from retrospective chart review and all participants came from tertiary referral centers, thus these results are prone to ascertainment bias and may not clearly reflect risks within the general AD population. As the focus of this study was to investigate factors that alter AD age at onset and clinical presentation, it also remains to be determined if, and to what extent, these factors confer increased risk of neurodegenerative disease. To do so will require future multicenter case-control studies that integrate the collection of these proposed novel factors across disease and healthy aging cohorts. As questions pertaining to early life learning differences and detailed health history are not routinely collected in healthy aging and dementia populations, we have proposed and piloted questionnaires specifically designed for collecting this type of data, to facilitate future standardized acquisition of this information, at scale, across centers. It also remains possible that the demographic differences we observed between EOAD and LOAD were influenced by changes in societal attitudes towards developmental differences, as well as survivor bias within our study population in those without autoimmune disease or seizure. With regards to societal bias, the practice of forced-right-handedness in the United States largely abated by 1945,(52) and thus should only pertain to the population age 75 and older within our study. To this end, in the MAC cohort, the amount of forced-right-handedness within our participants was no different between EOAD and LOAD participants (0.8%, n=6/750 vs. 0.3%, n=2/750), as such, we our hand preference results do not appear to be biased by changing attitudes in forced non-right-handedness. Conversely, widespread recognition of conditions like developmental dyslexia did not occur until the 1970’s,(53) therefore any bias in our collection regarding learning disability ascertainment would be expected to equally affect the EOAD and LOAD participants in this study, save the small percentage younger than 50 (6.4%, n=96/1500), nonetheless, we observed steady increases in learning disability prevelance with decreasing AD age at onset throught the standard EOAD/LOAD divide. With regards to the potential bias of survivor effects, without a healthy control comparison cohort, we cannot directly address the possibility of systematic drop out, due to premature death of individuals who suffered earlier in life from seizures or autoimmune disease within the LOAD cohort. However, the participants who possessed histories of active seizures are not subject to this survival bias concern, and as these cases demonstrate the same inverse linear association with age at onset as the remote seizures and autoimmune disease groups, it would strongly support that that the trends in autoimmunity and seizure reflect true risk factor profile differences between EOAD and LOAD. Another important limitation of this study is the lack of confirmed amyloid pathology across all participants. Given the nature of studies like the NACC and our own center’s database, a large fraction of participant data was collected prior to the advent and/or widespread use of AD biomarkers, limiting the ability to confirm AD pathology in those who did not present to autopsy. Despite these caveats, within the MAC cohort, nearly 1/3^rd^ of EOAD and 1/10_th_ of LOAD had confirmed amyloid positivity (either PET and/or autopsy proven), and within this group, all relative relationships between novel and typical factors remained (**Supplemental Table 4**), thus demonstrating clearly that the results presented here hold relevance for clinical and pathological AD presentations, alike.

In conclusion, in this study of over 10,000 clinically defined AD participants, we showed how hand preference, learning disability history, seizure, and autoimmune disease uniquely inform the age at onset, as well as phenotypical presentation of neurodegenerative disease, arguing for new branches of dementia epidemiology that focus on neurodevelopmental differences while broadening medical history beyond vascular disease. We also demonstrated how the interactions between these novel factors produced an exponential decrease in AD age at symptom onset, instigating our “two-hit” hypothesis of neurodegenerative disease risk, where neurodevelopmental factors interact with chronic neuroenvironmental insults to produce specific neurodegenerative disease patterns. Finally, given the ease of capturing these novel factors, along with their near-term ramifications for personalized disease prediction, intervention, and prevention, the results of this study underscore the acute need for acquiring detailed standardized histories of neurodevelopment and past medical conditions systematically across diverse healthy aging and dementia populations.

## Conflict of Interest Statement

**ZAM** reports no relevant disclosures. **RO** has been an associate editor for Alzheimer’s Research & Therapy, since 2018. **NGR** is partly funded by the David Eisenberg Mayo Clinic Professorship. **IEA** reports no relevant disclosures. **WS** reports no relevant disclosures. **LR** reports no relevant disclosures. **DJO** reports no relevant disclosures. **MGE** has received consultancy fees from Biogen. **PMB** is currently employed at Biogen but solely contributed to this work during fellowship at the UCSF Memory and Aging Center. **SS** reports receiving consulting fees from Acsel Health, Precision Xtract, and Techspert.io. **JY** reports grants from the National Institute on Aging during the conduct of the study, grants from National Institute on Aging, Rainwater Charitable Foundation, Transposon Therapeutics, Alector, and the US Department of Defense, and other support from the Mary Oakley Foundation and French Foundation outside the submitted work. **RSD** reports no disclosures. **WMVDF** is guest editor Alzheimer Research Therapy (series on SCD, 2018-2019); associate editor Alzheimer Research Therapy (2020 -). **PS** has received consultancy fees (paid to the institution) from AC Immune, Alkermes, Alnylam, Alzheon, Anavex, Biogen, Brainstorm Cell, Cortexyme, Denali, EIP, ImmunoBrain Checkpoint, GemVax, Genentech, Green Valley, Novartis, Novo Nordisk, PeopleBio, Renew LLC, Roche; he is PI of studies with AC Immune, CogRx, FUJI-film/Toyama, IONIS, UCB, Vivoryon; he is a part-time employee of Life Sciences Partners Amsterdam; he serves on the board of the Brain Research Center. **YALP** reports no relevant disclosures. **EEW** reports no relevant disclosures. **RR** reports no relevant disclosures. **DHG** reports no relevant disclosures. **JHK** reports no relevant disclosures. **HJR** reports no relevant disclosures. **KPR** reports no relevant disclosures. **LTG** reports no relevant disclosures. **WWS** reports no relevant disclosures. **VES** reports no relevant disclosures. **DCP** reports no relevant disclosures. **BLM** reports serving on the Cambridge National Institute for Health Research Biomedical Research Centre advisory committee and its subunit, the Biomedical Research Unit in Dementia; serving as a board member for the American Brain Foundation; serving on John Douglas French Alzheimer’s Foundation board of directors; serving on the Safely You board of directors; serving as scientific director for the Tau Consortium; serving as medical advisor for and receiving a grant from The Bluefield Project for Frontotemporal Dementia Research; serving as a consultant for Rainwater Charitable Foundation, Stanford Alzheimer’s Disease Research Center, Buck Institute SAB, Larry L. Hillblom Foundation, University of Texas Center for Brain Health, University of Washington Alzheimer’s Disease Research Center EAB, and Harvard University Alzheimer’s Disease Research Center EAB; receiving royalties from Guilford Press, Cambridge University Press, Johns Hopkins Press, and Oxford University Press; serving as editor for Neurocase; serving as section editor for Frontiers in Neurology; and receiving grants P30 AG062422, P01 AG019724, R01 AG057234, and T32 AG023481 from the NIH. **GDR** reports grants from the National Institutes of Health, Alzheimer’s Association, the American College of Radiology, Rainwater Charitable Foundation, Avid Radiopharmaceuticals, Eli Lilly, GE Healthcare, Life Molecular Imaging, and Genentech, as well as personal fees from Axon Neurosciences, Genentech, Johnson & Johnson, F. Hoffman–La Roche, and GE Healthcare outside the submitted work for service on scientific advisory boards (Axon Neurosciences, Eisai, Genentech, and F. Hoffman–La Roche) and a data safety monitoring board (Johnson & Johnson). **MLGT** reports no relevant disclosures.

## Author Contributions

All authors contributed to manuscript revision, read, and approved the submitted version. **ZAM** Conceptualized and designed the study, collected the data, performed formal analyses, was the primary author of the manuscript, and provided funding for the study. **RO** and **NGR** provided supervision over external cohort data collection from the Amsterdam and Mayo Clinic, Jacksonville, respectively, and interpretation of findings. **IEA** assisted with all aspects of the study design, methodology, interpretation of findings, performed formal analyses, and contributed to drafting and revision of the manuscript. **WS** and **LR** assisted with data collection. **MGE, PMB, SS, JY**, and **RSD** assisted with study design and interpretation of findings. **WMVDF, PS, YALP**, and **EEW** assisted with data collection from the Amsterdam cohort. **DJO, RR** assisted with data collection from the Mayo Clinic, Jacksonville cohort. **DHG, JHK, HJR, KPR, LTG**, and **WWS** assisted with data collection, study design and interpretation of findings. **VES** and **DCP** assisted with study design and interpretation of findings. **BLM, GDR**, and **MLGT** provided overall supervision, assisted with study design and interpretation of findings, and helped to provide funding for the study.

## Acknowledgements

We wish to express our gratitude to the participants and families who made this work possible. Additional inspiration for this work stems from an academic lineage beginning with Samuel Torrey Orton followed by Norman Geschwind and Al Galaburda, who codified the relationships between non-right-handedness, learning disability, immune disease, seizure, and more, into a single unified theory, as well as Frank Benson and Marsel Mesulam, who played pivotal roles in establishing the heterogeneity of neurodegenerative disease presentations.

The NACC database is funded by NIA/NIH Grant U01 AG016976. NACC data are contributed by the NIA-funded ADCs: P30 AG019610 (PI Eric Reiman, MD), P30 AG013846 (PI Neil Kowall, MD), P30 AG062428-01 (PI James Leverenz, MD) P50 AG008702 (PI Scott Small, MD), P50 AG025688 (PI Allan Levey, MD, PhD), P50 AG047266 (PI Todd Golde, MD, PhD), P30 AG010133 (PI Andrew Saykin, PsyD), P50 AG005146 (PI Marilyn Albert, PhD), P30 AG062421-01 (PI Bradley Hyman, MD, PhD), P30 AG062422-01 (PI Ronald Petersen, MD, PhD), P50 AG005138 (PI Mary Sano, PhD), P30 AG008051 (PI Thomas Wisniewski, MD), P30 AG013854 (PI Robert Vassar, PhD), P30 AG008017 (PI Jeffrey Kaye, MD), P30 AG010161 (PI David Bennett, MD), P50 AG047366 (PI Victor Henderson, MD, MS), P30 AG010129 (PI Charles DeCarli, MD), P50 AG016573 (PI Frank LaFerla, PhD), P30 AG062429-01(PI James Brewer, MD, PhD), P50 AG023501 (PI Bruce Miller, MD), P30 AG035982 (PI Russell Swerdlow, MD), P30 AG028383 (PI Linda Van Eldik, PhD), P30 AG053760 (PI Henry Paulson, MD, PhD), P30 AG010124 (PI John Trojanowski, MD, PhD), P50 AG005133 (PI Oscar Lopez, MD), P50 AG005142 (PI Helena Chui, MD), P30 AG012300 (PI Roger Rosenberg, MD), P30 AG049638 (PI Suzanne Craft, PhD), P50 AG005136 (PI Thomas Grabowski, MD), P30 AG062715-01 (PI Sanjay Asthana, MD, FRCP), P50 AG005681 (PI John Morris, MD), P50 AG047270 (PI Stephen Strittmatter, MD, PhD).

## Funding

This work was supported by National Institutes of Health grants (AG048291, AG053435, AG019724, AG062422, AG045611, AG062588, DC015544, and NS050915). Additional funds include the Hellman Research Scientist Award, the Arking Foundation for Frontotemporal Dementia, and the Jon and Gale Love fund.

## Notes

### Author Declarations

Ethics committees at the University of California San Francisco, Amsterdam University Medical Center, and Mayo Clinic, Jacksonville as well as all participating National Alzheimer's Coordinating Center sites approved the study of patients' clinical data.

