## Supplemental figures and tables for "Left-handedness, learning disability, autoimmune disease, and seizure history influence age at onset and phenotypical targeting of Alzheimer’s disease"

**Supplemental Table 1. Screen of Autoimmune Conditions.**

| Addison disease | Granulomatosis with Polyangiitis | Primary biliary cirrhosis |
| --- | --- | --- |
| Ankylosing spondylitis | Guillain-Barré syndrome | Psoriasis |
| Autoimmune hemolytic anemia | Hashimoto thyroiditis | Reactive arthritis |
| Autoimmune neutropenia | Inclusion body myositis | Rheumatoid arthritis |
| Behcet’s disease | Immune thrombocytopenic purpura | Sarcoidosis |
| Celiac disease | Lichen sclerosus | Sjögren’s syndrome |
| Chorea minor | Localized scleroderma | Systemic lupus erythematosus |
| Chronic lymphocitic colitis | Lupoid hepatitis | Systemic sclerosis |
| Chronic rheumatic heart disease | Multiple sclerosis | Transverse myelitis |
| Churg-Strauss Disease | Myasthenia gravis | Type 1 diabetes mellitus |
| Crohn’s disease | Pernicious anemia | Ulcerative colitis |
| Dermatomyositis | Polyarteritis nodosa | Vitiligo |
| Discoid lupus erythematosus | Polymyalgia rheumatic |  |
| Graves’ disease | Polymyositis |  |

Modified listing of autoimmune conditions screened from Rugbjerg et al.^17^

**Supplemental Table 2. MAC non-Amnestic AD vs. Remaining Amnestic EOAD & LOAD**

| **Group**  **Demographics** | **Amnestic EOAD**  **(n=555)** | **Amnestic LOAD**  **(n=661)** | **Amnestic EOAD**  **vs. Amnestic LOAD** | **non-Amnestic AD**  **(n=284)** | **non-Amnestic AD**  **vs. Amnestic EOAD**  **vs. Amnestic LOAD** |
| --- | --- | --- | --- | --- | --- |
| **Age at onset**  Average years ± Std | **55.9 ± 5.7** | **71.8 ± 4.1** | **<0.001** | **59.7 ± 7.9** | **<0.001** |
| **Age at diagnosis**  Average years ± Std | **60.6 ± 6.2** | **75.3 ± 4.3** | **<0.001** | **63.6 ± 7.9** | **<0.001** |
| **Sex**  % Male (n) | 43.6% (242/555) | 43.3% (286/661) | n.s. | 41.2% (117/284) | n.s. |
| **Typical Risk Factors** |  |  |  |  |  |
| **Education**  Average years ± Std | **15 ± 3.8**  **(540)** | **15 ± 3.9**  **(642)** | n.s. | **15.7 ± 2.8 (275)** | **0.02** |
| ***APOE*-ɛ4 carriers**  % with one or more ɛ4 alleles | **59.6%**  **(112/188)** | **65.3%**  **(81/124)** | n.s. | **44.2%**  **(65/147)** | **0.001** |
| **Hypertension**  % Hypertension (n) | **36.8% (204/555)** | **55.7% (368/661)** | **<0.001** | **35.2% (100/284)** | **<0.001** |
| **Hypercholesterolemia**  % Hypercholesterolemia (n) | **42.9% (238/555)** | **54.3% (359/661)** | **<0.001** | **45.1% (128/284)** | **<0.001** |
| **Diabetes**  % Diabetes Mellitus (n) | **9.2%**  **(51/555)** | **14.5%**  **(96/661)** | **0.004** | **6%**  **(17/284)** | **<0.001** |
| **Novel Factors** |  |  |  |  |  |
| **non-Right-Handed**  % non-right-handed (n) | **15%**  **(83/555)** | **7.7%**  **(51/661)** | **<0.001** | **15.8%**  **(45/284)** | **<0.001** |
| **Learning Disability**  % Learning Disability (n) | **5.8%**  **(32/555)** | **0.8%**  **(5/661)** | **<0.001** | **22.5%**  **(64/284)** | **<0.001** |
| **Remote Seizure**  % Seizure (n) | **4%**  **(22/555)** | **1.2%**  **(8/661)** | **0.002** | **2.1%**  **(6/284)** | **0.007** |
| **Active Seizure**  % Seizure (n) | **7.7%**  **(43/555)** | **3.5%**  **(23/661)** | **0.001** | **3.5%**  **(10/284)** | **0.001** |
| **Autoimmune Disease**  % Autoimmunity (n) | **24.3% (135/555)** | **19.7% (130/661)** | **0.05** | 25.7%  (73/284) | n.s. |

AD = Alzheimer’s disease; EOAD = early-onset Alzheimer’s disease; lvPPA = logopenic variant primary progressive aphasia; n.s. = not significant; PCA = posterior cortical atrophy; Std = standard deviation.

Compared to amnestic LOAD, amnestic EOAD has less hypertension, hypercholesterolemia, diabetes mellitus and more non-right-handedness, learning disability, seizure, and autoimmune disease.

**Supplemental Table 3. non-Amnestic AD Broken into lvPPA & PCA Subgroups**

| **Group**  **Demographics** | **non-Amnestic** **(n=284)** | **lvPPA**  **(n=171)** | **PCA**  **(n=113)** | **lvPPA vs. PCA** |
| --- | --- | --- | --- | --- |
| **Age at onset**  Average years ± Std | 59.7 ± 7.9 | **60.7 ± 8.3** | **58.1 ± 7.1** | **0.007** |
| **Age at diagnosis**  Average years ± Std | 63.6 ± 7.9 | **64.5 ± 8.1** | **62.1 ± 7.2** | **0.01** |
| **Sex**  % Male (n) | 41.2% (117/284) | 45.6%  (78/171) | 34.5%  (39/113) | n.s. |
| **Typical Risk Factors** |  |  |  |  |
| **Education**  Average years ± Std | 15.7 ± 2.8 (275) | 15.7 ± 2.8 (164) | 15.8 ± 3  (111) | n.s. |
| ***APOE*-ɛ4 carriers**  % with one or more ɛ4 alleles | 44.2%  (65/147) | 43.8%  (39/89) | 44.8%  (26/58) | n.s. |
| **Hypertension**  % Hypertension (n) | 35.2% (100/284) | 37.4%  (64/171) | 31.9%  (36/113) | n.s. |
| **Hypercholesterolemia**  % Hypercholesterolemia (n) | 45.1% (128/284) | 46.8%  (80/171) | 42.5%  (48/113) | n.s. |
| **Diabetes**  % Diabetes Mellitus (n) | 6%  (17/284) | **8.8%**  **(15/171)** | **1.8%**  **(2/113)** | **0.01** |
| **Novel Factors** |  |  |  |  |
| **non-Right-Handed**  % non-right-handed (n) | 15.8%  (45/284) | 12.9%  (22/171) | 20.4%  (23/113) | n.s. |
| **Learning Disability**  % Learning Disability (n) | 22.5%  (64/284) | 21.1%  (36/171) | 24.8%  (28/113) | n.s. |
| **Remote Seizure**  % Seizure (n) | 2.1%  (6/284) | 1.8%  (3/171) | 2.7%  (3/113) | n.s. |
| **Active Seizure**  % Seizure (n) | 3.5%  (10/284) | **1.8%**  **(3/171)** | **6.2%**  **(7/113)** | **0.047** |
| **Autoimmune Disease**  % Autoimmunity (n) | 25.7%  (73/284) | **17.5%**  **(30/171)** | **38.1%**  **(43/113)** | **<0.001** |

AD = Alzheimer’s disease; EOAD = early-onset Alzheimer’s disease; lvPPA = logopenic variant primary progressive aphasia; n.s. = not significant; PCA = posterior cortical atrophy; Std = standard deviation.

Compared to lvPPA, the PCA group was 2.6 years younger at age of onset, 2.4 years younger at age of diagnosis, has less diabetes, more active seizure, and more autoimmune disease.

**Supplemental Table 4. Demographics of Amyloid Confirmed vs. Unknown MAC AD**

|  | **non-Amnestic AD** | | **Amnestic EOAD** | | **Amnestic LOAD** | |
| --- | --- | --- | --- | --- | --- | --- |
| **Group** | **Amyloid Confirmed** | **Amyloid Unknown** | **Amyloid Confirmed** | **Amyloid Unknown** | **Amyloid Confirmed** | **Amyloid Unknown** |
| **Demographics** | **(n=114)** | **(n=170)** | **(n=118)** | **(n=437)** | **(n=41)** | **(n=620)** |
| **Age at onset**  Average years ± Std | **57.8 ± 7.0**** | **60.9 ± 8.2**** | **53.8 ± 5.4**** | **56.5 ± 5.6**** | 71.6 ± 3.9 | 71.9 ± 4.1 |
| **Age at diagnosis**  Average years ± Std | **61.7 ± 7.1**** | **64.9 ± 8.0**** | **58.2 ± 5.8**** | **61.2 ± 6.1**** | 74.8 ± 3.4 | 75.3 ± 4.3 |
| **Sex**  % Male (n) | 38.6% (44/114) | 42.9% (73/170) | **55.9%** (66/118)** | **40.3%** (176/437)** | **61.0%***  **(25/41)** | **42.1%***  **(261/620)** |
| **Typical Risk Factors** |  |  |  |  |  |  |
| **Education**  Average years ± Std | **16.4 ± 2.4** (111)** | **15.3 ± 3.0** (164)** | **16.3 ± 2.6** (115)** | **14.7 ± 4.0****  **(425)** | **16.6 ± 3.2* (41)** | **14.9 ± 3.9* (601)** |
| ***APOE*-ɛ4 carriers**  % with one or more ɛ4 alleles | 43.1%  (44/102) | 46.7%  (21/45) | 59.0%  (59/100) | 60.2%  (53/88) | 69.0%  (20/29) | 64.9%  (61/94) |
| **Hypertension**  % Hypertension (n) | **25.4%* (29/114)** | **41.8%* (71/170)** | **23.7%** (28/118)** | **40.3%** (176/437)** | 46.3%  (19/41) | 56.3% (349/620) |
| **Hypercholesterolemia**  % Hypercholesterolemia (n) | **52.6%* (60/114)** | **40.0%* (68/170)** | 38.1% (45/118) | 44.2%  (193/437) | 61.0% (25/41) | 53.9% (334/620) |
| **Diabetes**  % Diabetes Mellitus (n) | 5.3%  (6/114) | 6.5%  (11/170) | **4.2%* (5/118)** | **10.5%* (46/437)** | 22.0%  (9/41) | 14.0%  (87/620) |
| **Novel Factors** |  |  |  |  |  |  |
| **non-Right-Handed**  % non-right-handed (n) | 16.7% (19/114) | 15.3%  (26/170) | 15.2% (18/118) | 14.9% (65/437) | 9.8%  (4/41) | 7.6%  (47/620) |
| **Learning Disability**  % Learning Disability (n) | **30.7%***  **(35/114)** | **17.1%* (29/170)** | 6.8%  (8/118) | 5.5%  (24/437) | 0.0%  (0/41) | 0.8%  (5/620) |
| **Remote Seizure**  % Seizure (n) | 1.8 % (2/114) | 2.4%  (4/170) | 2.5% (3/118) | 4.3%  (19/437) | 0.0%  (0/41) | 1.3%  (8/620) |
| **Active Seizure**  % Seizure (n) | 3.5% (4/114) | 3.5%  (6/170) | 8.5% (10/118) | 7.6%  (33/437) | 2.4%  (1/41) | 3.5%  (22/620) |
| **Autoimmune Disease**  % Autoimmunity (n) | 25.4% (29/114) | 25.8% (44/170) | 18.6% (22/118) | 25.9% (113/437) | 19.5%  (8/41) | 19.7% (122/620) |

* < 0.05; ** < 0.001

AD = Alzheimer’s disease; Std = standard deviation.

**Supplemental Table 5. MAC and NACC & AUMC Broken into AD Age at Onset Quintiles**

|  | **MAC 1,500 AD** | | | | | **NACC & AUMC AD** | | | | |
| --- | --- | --- | --- | --- | --- | --- | --- | --- | --- | --- |
| **Group** | **1** | **2** | **3** | **4** | **5** | **1** | **2** | **3** | **4** | **5** |
| **Demographics** | **(n=300)** | **(n=300)** | **(n=300)** | **(n=300)** | **(n=300)** | **(n=1772)** | **(n=1772)** | **(n=1772)** | **(n=1772)** | **(n=1771)** |
| **Age at onset**  Average years ± Std | 50.2 ± 3.1 | 57.7 ± 2.2 | 64.6 ± 1.8 | 69.9 ± 1.6 | 75.9 ± 2.1 | 55.3 ± 5.2 | 65.9 ± 2.1 | 71.6 ± 1.4 | 76.2 ± 1.4 | 82.9 ± 3.6 |
| **Age at diagnosis**  Average years ± Std | 54.9 ± 3.9 | 62 ± 3.6 | 68.7 ± 3 | 73.5 ± 2.8 | 78.9 ± 2.5 | 60.6 ± 6.3 | 71.3 ± 4.2 | 76.6 ± 3.5 | 80.6 ± 3.1 | 86.4 ± 4.1 |
| **Sex**  % Male (n) | 40% (120/300) | 42.7% (128/300) | 46.7% (140/300) | 43.7% (131/300) | 42% (126/300) | 47.7% (846/1772) | 44.2% (783/1772) | 44.5% (789/1772) | 41.7% (739/1772) | 39.9% (707/1771) |
| **Typical Risk Factors** |  |  |  |  |  |  |  |  |  |  |
| **Education**  Average years ± Std | 15 ± 3.2 (295) | 15.3 ± 3.7 (289) | 15.3 ± 4.1 (289) | 15.2 ± 3.8 (291) | 15 ± 3.6 (293) | 14.3 ± 3.7 (1756) | 14.1 ± 3.7 (1764) | 13.8 ± 4 (1763) | 13.7 ± 3.9 (1753) | 13.3 ± 4.1 (1761) |
| ***APOE*-ɛ4 carriers**  % w/ɛ4 alleles | 52.2%  (70/134) | 52.1% (62/119) | 60.5%  (49/81) | 61.9%  (39/63) | 61.3%  (38/62) | 58.3%  (792/1357) | 71.4%  (919/1288) | 64.6%  (820/1269) | 56.5%  (712/1261) | 42.7%  (500/1172) |
| **Hypertension**  % HTN (n) | 27.7% (83/300) | 37.7% (113/300) | 47.3% (142/300) | 55.3% (166/300) | 56% (168/300) | 34.8% (615/1768) | 50.8% (897/1765) | 56.5% (997/1766) | 60.1% (1064/1769) | 62% (1096/1767) |
| **Hypercholesterolemia** % HCL (n) | 34% (102/300) | 45.3% (136/300) | 54.3% (163/300) | 55% (165/300) | 53% (159/300) | 41.1% (722/1757) | 55.3% (971/1757) | 53.6% (941/1756) | 52% (910/1750) | 46.2% (805/1741) |
| **Diabetes**  % DM (n) | 6% (18/300) | 8.7% (26/300) | 13.7% (41/300) | 13% (39/300) | 13.3% (40/300) | 9.2% (162/1764) | 13.6% (240/1770) | 15.9% (282/1770) | 14.1% (249/1768) | 13.1% (231/1766) |
| **Novel Factors** |  |  |  |  |  |  |  |  |  |  |
| **non-Right-Handed**  % nRH (n) | 13.7% (41/300) | 15.7% (47/300) | 12.7% (38/300) | 11% (33/300) | 6.7% (20/300) | 9.8% (173/1764) | 7.7% (136/1770) | 9.7% (171/1768) | 7.6% (134/1768) | 6.8% (120/1770) |
| **Learning Disability**  % LD (n) | 13% (39/300) | 11.3% (34/300) | 6%  (18/300) | 3%  (9/300) | 0.3%  (1/300) |  |  |  |  |  |
| **Remote Seizure**  % Seizure (n) | 4.3% (13/300) | 4.3% (13/300) | 0.7%  (2/300) | 1.7%  (5/300) | 1%  (3/300) | 2.5% (43/1712) | 1.7% (30/1735) | 1.6% (27/1737) | 1.1% (19/1755) | 0.5% (8/1753) |
| **Active Seizure**  % Seizure (n) | 8% (24/300) | 7.3% (22/300) | 4.3% (13/300) | 4%  (12/300) | 1.7%  (5/300) | 2.7% (47/1759) | 1.7% (30/1765) | 1.4% (25/1762) | 0.7% (13/1768) | 0.7% (12/1765) |
| **Autoimmune Disease**  % Autoimmunity (n) | 25.3% (76/300) | 24.3% (73/300) | 24% (72/300) | 21.7% (65/300) | 17.3% (52/300) |  |  |  |  |  |
| **non-Amnestic** **AD**  % non-Amnestic AD (n) | 25.7% (77/300) | 30.7% (92/300) | 18.7% (56/300) | 15% (45/300) | 4.7% (14/300) | 4.8% (85/1772) | 1.2% (22/1772) | 1% (17/1772) | 0.5% (8/1772) | 0.1% (2/1771) |

AD = Alzheimer’s disease; DM = diabetes mellitus; HCL = hypercholesterolemia; HTN = hypertension; LD = learning disability; nRH = non-right-handed; Std = standard deviation.

Within each cohort, placing each subject in order of age at first symptom onset, we split each cohort evenly into quintiles, 300 subjects within each MAC quintile and 1772 within each NACC & AUMC quintile. The average age of each quintile was obtained. Similarly, the percentage within each quintile of the various factors was calculated. We plotted out each quintile based on their average age at onset by the percentage factor of interest (**Figure 4**). The novel factors all possess inverse linear associations with age at onset, while the typical risk factors appear to be best described by quadratic relationships with age at onset.

**Supplemental Table 6. Interactions Between Novel Factors in the MAC Cohort**

| **Group** | **RH** | **nRH** | **nonLD** | **LD** | **No Seizures** | **Remote Seizures** | **Active Seizures** | **No Autoimmune** | **Autoimmune** |
| --- | --- | --- | --- | --- | --- | --- | --- | --- | --- |
| **Demographics** | **(n=1321)** | **(n=179)** | **(n=1399)** | **(n=101)** | **(n=1400)** | **(n=36)** | **(n=76)** | **(n=1162)** | **(n=338)** |
| **Age at onset**  Average years ± Std | **64 ± 9.3**** | **61.4 ± 8.7**** | **64.2 ± 9.1**** | **56.2 ± 7.5**** | **64 ± 9.2**** | **59.3 ± 9.1**** | **59.6 ± 8.7**** | **63.9 ± 9.3*** | **62.7 ± 9*** |
| **Age at diagnosis**  Average years ± Std | **67.9 ± 9.1*** | **65.6 ± 8.6*** | **68.1 ± 8.9**** | **60.4 ± 7.6**** | **67.9 ± 9**** | **63.5 ± 8.9**** | **63.7 ± 8.7**** | **67.9 ± 9.1*** | **66.7 ± 8.8*** |
| **Sex**  % Male (n) | **41.9%* (553/1321)** | **51.4%* (92/179)** | 43.4% (607/1399) | 37.6% (38/101) | 42.9% (600/1400) | 47.2% (17/36) | 46.1% (35/76) | **48%****  **(558/1162)** | **25.7%****  **(87/338)** |
| **Typical Risk Factors** |  |  |  |  |  |  |  |  |  |
| **Education**  Average years ± Std | **15.1 ± 3.8* (1283)** | **15.9 ± 2.9* (174)** | 15.1 ± 3.7 (1360) | 15.6 ± 3  (97) | 15.2 ± 3.7 (1360) | 14.4 ± 4.6 (35) | 15.2 ± 2.9 (73) | 15.2 ± 3.7 (1128) | 15.2 ± 3.5 (329) |
| ***APOE*-ɛ4 carriers**  % w/ɛ4 alleles | 57.7%  (228/395) | 46.9%  (30/64) | **58.5%***  **(234/400)** | **40.7%***  **(24/59)** | 56.7%  (237/418) | 33.3%  (5/15) | 62.1%  (18/29) | 57.7%  (211/366) | 50.5%  (47/93) |
| **Hypertension**  % HTN (n) | 45.2% (597/1321) | 41.9% (75/179) | **45.9%* (642/1399)** | **29.7%* (30/101)** | 45.4% (635/1400) | 44.4% (16/36) | 35.5% (27/76) | **46.7%* (543/1162)** | **38.2%* (129/338)** |
| **Hypercholesterolemia**  % HCL (n) | 49.1% (648/1321) | 43% (77/179) | **49.2%* (688/1399)** | **36.6%* (37/101)** | 48.4% (678/1400) | 52.8% (19/36) | 44.7% (34/76) | 48.9% (568/1162) | 46.4% (157/338) |
| **Diabetes**  % DM (n) | **11.7%* (154/1321)** | **5.6%***  **(10/179)** | 10.9% (153/1399) | 10.9% (11/101) | 11% (154/1400) | 11.1%  (4/36) | 7.9%  (6/76) | 10.8% (125/1162) | 11.5%  (39/338) |
| **Novel Factors** |  |  |  |  |  |  |  |  |  |
| **non-Right-Handed**  % nRH (n) |  |  | **10.9%** (153/1399)** | **25.7%** (26/101)** | 12% (168/1400) | 11.1%  (4/36) | 10.5%  (8/76) | 12.3% (143/1162) | 10.7%  (36/338) |
| **Learning Disability**  % LD (n) | **5.7%** (75/1321)** | **14.5%** (26/179)** |  |  | 6.6% (93/1400) | 2.8%  (1/36) | 9.2%  (7/76) | 6.9%  (80/1162) | 6.2%  (21/338) |
| **Remote Seizure**  % Seizure (n) | 2.4% (32/1321) | 2.2%  (4/179) | 2.5% (35/1399) | 1%  (1/101) |  |  |  | 2.4%  (28/1162) | 2.4%  (8/338) |
| **Active Seizure**  % Seizure (n) | 5.1% (68/1321) | 4.5%  (8/179) | 4.9% (69/1399) | 6.9%  (7/101) |  |  |  | **4.3%***  **(50/1162)** | **7.7%***  **(26/338)** |
| **Autoimmune Disease**  % Autoimmunity (n) | 22.9% (302/1321) | 20.1% (36/179) | 22.7% (317/1399) | 20.8% (21/101) | **22%* (308/1400)** | **22.2%***  **(8/36)** | **34.2%* (26/76)** |  |  |
| **non-Amnestic** **AD**  % non-Amnestic AD (n) | **18.1%* (239/1321)** | **25.1%* (45/179)** | **15.7%** (220/1399)** | **63.4%** (64/101)** | 19.2% (269/1400) | 16.7%  (6/36) | 13.2% (10/76) | 18.2% (211/1162) | 21.6%  (73/338) |

* < 0.05; ** < 0.001

AD = Alzheimer’s disease; DM = diabetes mellitus; HCL = hypercholesterolemia; HTN = hypertension; LD = learning disability; nonLD = non-learning disability; nRH = non-right-handed; RH = right-handed; Std = standard deviation.

**Supplemental Figure 1. Interactions between *APOE*-ɛ4 and Novel Factors in non-Amnestic AD and Remaining EOAD & LOAD cohorts**

A. **non-Amnestic AD with Novel Factors Split by *APOE*-ɛ4 Status**

***APOE-*ɛ4 negative**

***APOE-*ɛ4 positive**

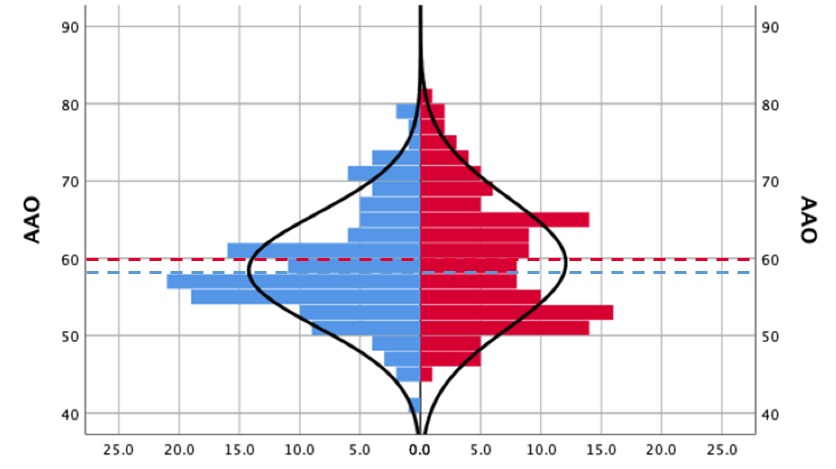

B. **EOAD & LOAD without non-Amnestic AD, Novel Factors Split by *APOE*-ɛ4 Status**

***APOE-*ɛ4 negative**

***APOE-*ɛ4 positive**

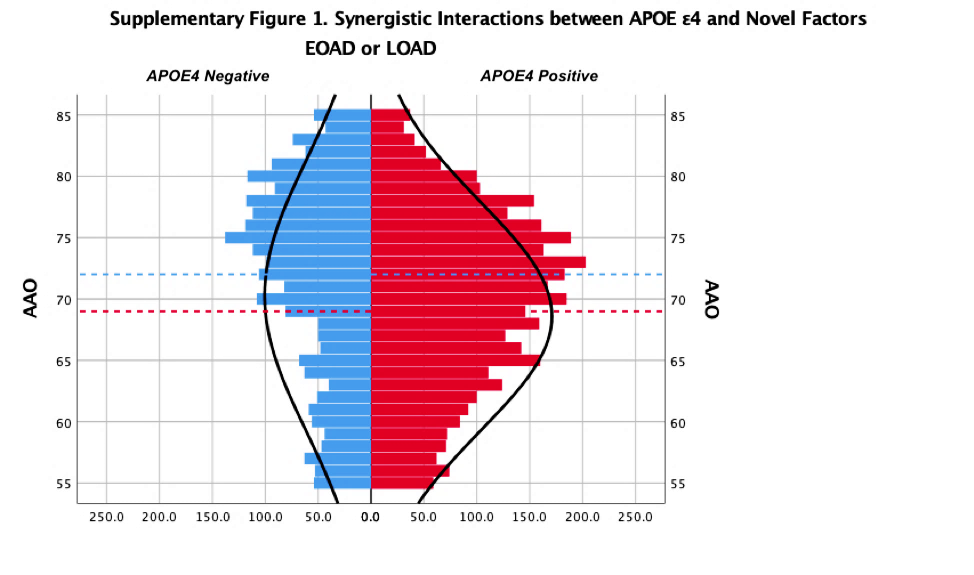

***APOE-*ɛ4 negative**

***APOE-*ɛ4 positive**

A. From combined MAC and NACC & AUMC cohorts, we plotted histograms of all non-amnestic AD who possessed novel factors (nRH and seizure), dichotomized by APOE-ɛ4 status (on the left, in blue for those who were *APOE-ɛ4* negative, and on the right, in red for *APOE-ɛ4* positive cases). Mean AAO for novel factors within the *APOE-ɛ4* negative group was 58±7.3 years (blue dashed line) and for the *APOE-ɛ4* positive group 60±8.4 years (red dashed line). These means were not statistically different from each other.

B. Plotting histograms of all remaining EOAD & LOAD who possessed novel factors (nRH and seizure) dichotomized by *APOE-ɛ4* status the means were statistically different from each other (p=0.013), 72±10.9 years for the APOE-ɛ4 negative group and 69±9.3for and for the APOE-ɛ4positive group.

**Supplemental Figure 2. Burden of Factors on Age at Onset within MAC and NACC & AUMC**
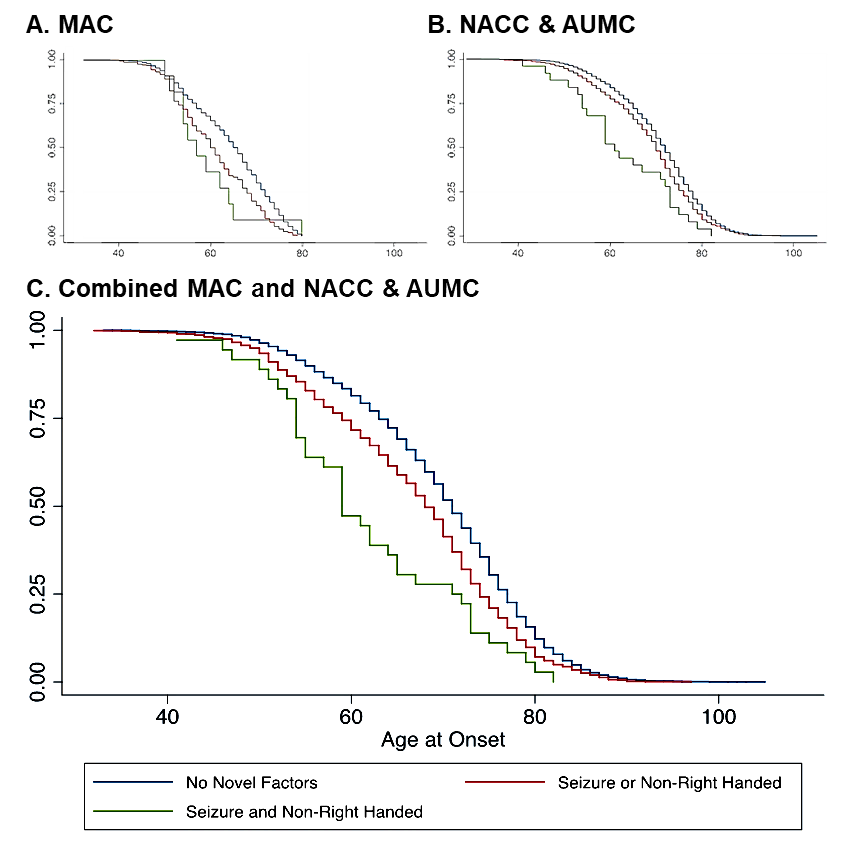

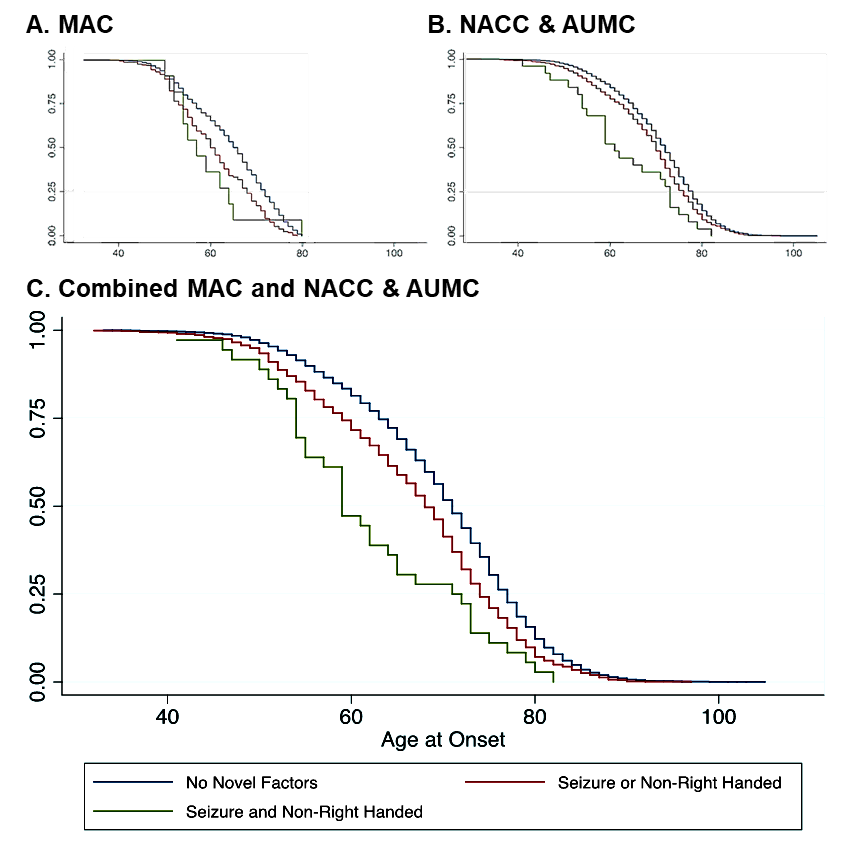

Plotting individuals stratified by numbers of novel factors (non-right-handedness and seizure) versus the age at which they developed first symptoms produced three distinct Kaplan-Meier curves.

In the MAC cohort (A) the blue line (n=1232) encompasses all participants who lacked any novel factor. The red line (n=257) included only those who had one novel factor (either non-right-handedness or seizure). The green line (n=11) comprised those individuals who possessed both novel factors (non-right-handedness and seizure). Using ANOVA with a Scheffe correction for multiple comparisons: No Novel factors vs. one Novel factor (blue vs. red) *p*<0.001; No Novel factors vs. two Novel factors (blue vs. green) *p*=0.047; One Novel factor vs. two Novel factors (red vs. green) *p*=0.102.

In the NACC & AUMC cohort (B) the blue line (n=7842) encompasses all participants who lacked any novel factor. The red line (n=933) included only those who had one novel factor (either non-right-handedness or seizure). The green line (n=25) comprised those individuals who possessed both novel factors (non-right-handedness and seizure). Using ANOVA with a Scheffe correction for multiple comparisons: No Novel factors vs. one Novel factor (blue vs. red) *p*<0.001; No Novel factors vs. two Novel factors (blue vs. green) *p*<0.001; One Novel factor vs. two Novel factors (red vs. green) *p*=0.006
